# Modeling dynamic disease-behavior feedbacks for improved epidemic prediction and response

**DOI:** 10.1101/2024.11.16.24317352

**Authors:** Hongru Du, Matthew Zahn, Sara Loo, Tijs Alleman, Shaun Truelove, Bryan Patenaude, Lauren Gardner, Nicholas Papageorge, Alison Hill

## Abstract

Human behavior significantly influences infectious disease transmission, yet traditional models often overlook this factor, limiting predictions of disease and the associated socioeconomic impacts. We introduce a feedback-informed epidemiological model that integrates economic decision-making with infectious disease dynamics. Individuals weigh costs and benefits, then choose behaviors that influence their risk of infection and disease progression, thereby shaping population-level dynamics. Applying this model to a scenario based on the early COVID-19 pandemic, we examine decisions to abstain from work to mitigate infection risk. Our findings reveal that feedback between disease and behavior notably affect infection rates and overall welfare, especially when accounting for individual economic and health vulnerabilities, which are often in tension. We evaluate counterfactual policies, including labor restrictions and cash transfers, illustrating how our framework can simultaneously address epidemiological, economic, and equity-related questions. This flexible and extendable modeling framework offers a powerful tool for assessing infectious disease interventions.

## Introduction

Infectious disease transmission is driven by human behavior, which brings people in contact with the pathogens we host. For most of human history, behavioral modifications to reduce transmission, such as quarantine and isolation, were the main methods of infection control. Even today, the choice to wash hands, get tested, vaccinated or take medication drives individual and collective risk for many diseases. During the early years of the COVID-19 pandemic, policies to induce widespread behavior changes such as business/school closures, stay-at-home orders, and travel bans were common. While these interventions dramatically reduced disease spread and healthcare burden for some time, they also caused substantial disruptions to well-being. Thus, a recurring question facing policy-makers has been “How do we reduce disease burden while simultaneously mitigating the social and economic costs of doing so?”

Disease transmission models are powerful tools for informing control policy, as evidenced by their widespread use for vaccine-preventable or drug-treatable infections such as COVID-19, HIV, influenza, measles, and malaria (e.g. Cramer et al., 2022; Loo et al., 2024; Smith et al., 2012; Eaton et al., 2012; Anderson et al., 1991; Keeling and Grenfell, 1997; Winter et al., 2022). Historically, these models track changes in the portion of a population at risk of infection and in different stages of disease progression (e.g., the SIR model Kermack and McKendrick, 1927). Transitions between stages are determined by composite parameter values informed by epidemiological observations—such as the probability of transmission per time period for a given population density or the average duration of infectiousness—which obscures the specific impact of human behavior or the generative process governing it. During COVID-19 and to some extent before, data such as contact surveys (Feehan and Mahmud, 2021; Wallinga et al., 2006), mobility metrics (Nouvellet et al., 2021; Wesolowski et al., 2016), or real-time vaccination tracking (Salomon et al., 2021) allowed models to include modifications to parameter values based on these data-derived correlates of behavior, often at high temporal or spatial resolution (Gog et al., 2014) or stratified by known risk factors, for example, age (Prem et al., 2017; Mistry et al., 2021). However, these approaches abstract from the mechanisms behind individual-level decision making and thus fail to capture the dynamic trade-offs between health and other aspects of well-being that individuals face as disease burden and control strategy evolve. To adequately capture these trade-offs, we need modeling frameworks that account for the complex interactions between disease propagation and the behaviors that drive it, including feedback loops (whereby behavior change leads to changes in disease dynamics that in turn lead to further shifts in behavior), externalities (whereby individual choices have an impact on others in society), and heterogeneity in decision making (whereby individuals may face different trade-offs depending on their health or economic vulnerability). Absent models that capture these complexities, it is difficult to generate reliable projections of disease transmission or to evaluate the welfare consequences—including economic costs—of prospective public health policies.

Health economists have long integrated infectious disease models in cost-benefit and cost-effectiveness analyses to guide public health policy (Fisman and Tuite, 2011; Nosyk et al., 2015). However, these approaches often rely on simple models of disease spread that rarely consider feedback among disease prevalence, individual behavior, and public policy (Vandepitte et al., 2021). Projected health outcomes are typically converted to disability- or quality-adjusted life years(Anand and Hanson, 1997), but these metrics do not encompass an individual’s overall well-being (Williams, 1999; Nord, 2016). Furthermore, the economic analyses accompanying these studies tend to focus narrowly on direct medical costs and certain indirect costs such as lower productivity, neglecting costs stemming from behavioral changes, income loss, and the broader disutility of policy constraints. This omission can lead to underestimates of the full cost of the disease or policies to curb it, along with inaccurate predictions about behavior and thus disease spread.

The study of how individuals weigh trade-offs to make decisions in a variety of circumstances—including infectious disease outbreaks—is a substantial part of research in economics. Prior work has integrated disease dynamics into models of human behavior related to labor supply, consumption, and risky behaviors (see e.g., Kremer, 1996; Auld, 2003; Chan et al., 2015; Papageorge, 2016; Greenwood et al., 2019; Keogh-Brown et al., 2010). With a primary goal of better understanding human behavior, these studies have placed less emphasis on the epidemiological components, potentially leading to misspecifications of how diseases are contracted, transmitted, or progress. Despite capturing how behavior endogenously responds to prevailing disease conditions, these models thus tend to be ill-equipped to forecast disease dynamics, which in turn can lead to inaccurate forecasts of behavioral responses and evaluations of intervention policies.

To address these challenges, a growing body of research in behavioral epidemiology and economic epidemiology has begun developing integrated frameworks of disease spread and human behavior(e.g., Gersovitz and Hammer, 2004; Manfredi and D’Onofrio, 2013; Funk et al., 2010; Fenichel, 2013; Funk et al., 2015; Bedson et al., 2021; Darden et al., 2022; Weston et al., 2018; Verelst et al., 2016; Perrings et al., 2014; Dangerfield et al., 2022). Prior models have included reasonable approximations to both pathogen transmission and behavior. For example, traditional infectious disease models have been extended to include heuristic functions for changes in contact rates with disease burden (e.g., Capasso and Serio, 1978; Morin et al., 2013; Glaubitz and Fu, 2020; Arthur et al., 2021), to model the spread or “imitation” of behaviors contemporaneously with infection (see e.g., Tanaka et al., 2002; Funk et al., 2009), or to consider behavior as a game-theory problem where disease levels are static on the timescale of decision making and large groups of the population collapse into a small number of “players” all making the same sets of decisions(e.g., Chen, 2004; Reluga, 2010; Chang et al., 2020; Saad-Roy and Traulsen, 2023; Martcheva et al., 2021).

Inspired by the needs of policymakers during the COVID-19 pandemic, new approaches to model behavior and disease spread have emerged. One leverages macroeconomic models assuming non-infected individuals supply labor thereby contribute to aggregate output (Pichler et al., 2020; Acemoglu et al., 2021; Fabbri et al., 2021; Pichler et al., 2022; Haw et al., 2022; Pangallo et al., 2023; Dobson et al., 2023; Alleman and Baetens, 2024). These frameworks forecast policy-relevant indicators such as unemployment and gross domestic product, but, without a formal model of individual-level decision making (or by assuming imitation), cannot fully capture the feedbacks and trade-offs that influence economically-relevant behavior and therefore do not adequately capture the welfare consequences of policy. Approaches that do directly model how individuals make decisions often employ fixed decision rules, sometimes informed by data, to predict how behavior will respond to prevailing disease conditions (Zanette and Risau-Gusmán, 2008; Perisic and Bauch, 2009b,a; McAdams et al., 2019; Khanjanianpak et al., 2022; Morsky et al., 2023). Given fixed decision rules, this approach is not designed to capture how individuals re-optimize under counterfactual policies or disease scenarios.

A handful of prior papers have incorporated formal models of behavior where decisions are made to optimize a measure of well-being or utility with potentially incomplete information, and can thus project how behavior endogenously responds to changing disease and policy conditions (Fenichel et al., 2011; Eichenbaum et al., 2021; Ash et al., 2022; Brotherhood et al., 2023). However, these studies have two main limitations. First, some have ignored individual heterogeneity in vulnerability to disease (e.g., preexisting conditions) or economic hardship (e.g., low income), instead differentiating individuals *only* by their infection state (Fenichel et al., 2011; Eichenbaum et al., 2021; Ash et al., 2022). Capturing population heterogeneity is critical not only for quantifying the distributional benefits and burdens of different policy interventions, but also for accurately predicting population-level disease spread, as concentration of infection in risk groups promotes persistence despite control efforts. A second limitation of this body of work is the use of non-standard or inflexible approaches to describing infection spread (Chan et al., 2015; Greenwood et al., 2019; Brotherhood et al., 2023; Aguirregabiria et al., 2021). For example Brotherhood et al. (2023) capture important margins of individual heterogeneity in their model of behavior, but make limiting assumptions in their epidemiological model (e.g., random mixing, no group stratification, calibrated disease dynamics).

In this paper, we present a parsimonious integrated framework that draws from economics and epidemiology to capture the feedback between human behavior and disease spread. From economics, we draw on models of dynamic decision making at the individual level, which responds to and provides inputs to an epidemiological model of population disease spread. Heterogeneous agents compare costs and benefits of different behavioral choices and assess the associated disease risk, considering current and future payoffs specific to their individual characteristics. These decisions affect individuals’ risks of acquiring, transmitting, and developing severe outcomes as a result of infection, which in turn drives population-level disease dynamics and thus future decisions. These features allow us to project disease outcomes and, importantly, evaluate the macro-level outputs of counterfactual policies in a manner that accounts for endogenous behavioral responses and trade-offs between health and economic well-being (that have heterogeneous effects on different sub-populations). As an example application, we use our model to evaluate disease-decision feedbacks during the early months of the COVID-19 pandemic, when the decision to participate in paid, in-person work was a major determinant of disease risk. We include two crucial dimensions of heterogeneity—health and economic vulnerability. Comparing the impacts of stylized policy options representing mandatory, incentivized/compensated, and voluntary work abstention, we find that accounting for disease-behavior feedback has a significant impact on the relative health and economic impacts of policies, and on how inequities between risk groups are exacerbated or alleviated by disease control measures. Importantly, we show that it is plausible to both reduce disease burden and also increase labor supply compared to the *status quo*, generally seen as an elusive goal given tensions between health and economic well-being during a pandemic.

## Results

### Development of a feedback-informed epidemiological model

In this paper, we present a dynamic Feedback-Informed Epidemiological Model (FIEM) that integrates infectious disease dynamics with individual behavior. Our framework classifies individuals based on their time-varying infection status (e.g., susceptible, infectious) and decision status (e.g., choice to work or engage in social distancing), as well as by a set of other state variables that may be fixed or time-dependent (e.g., demographics, health vulnerability, socioeconomic profile). The two core components of the dynamic mathematical model—the risk-stratified model of disease transmission and the individual-level model of decision-making—determine how individuals’ infection and decision states evolve over time (Figure 1). We designed FIEM to be flexible, allowing the disease and decision models to be extended in many possible directions, such as adding more infection states (e.g., asymptomatic, mild-symptomatic), incorporating additional decision sets (e.g., compliance with mask mandates, willingness to take vaccine, allocating time between work and leisure), or specifying new state variables to further differentiate individuals. Together these characteristics make FIEM a powerful and flexible tool for policy analysis; generating predictions about disease spread and economic consequences that capture endogenous individual decision making, and allowing for analysis of the impact of policy interventions across different types of individuals in the population. The components of the model are summarized here and detailed in the Methods.

**Figure 1:**
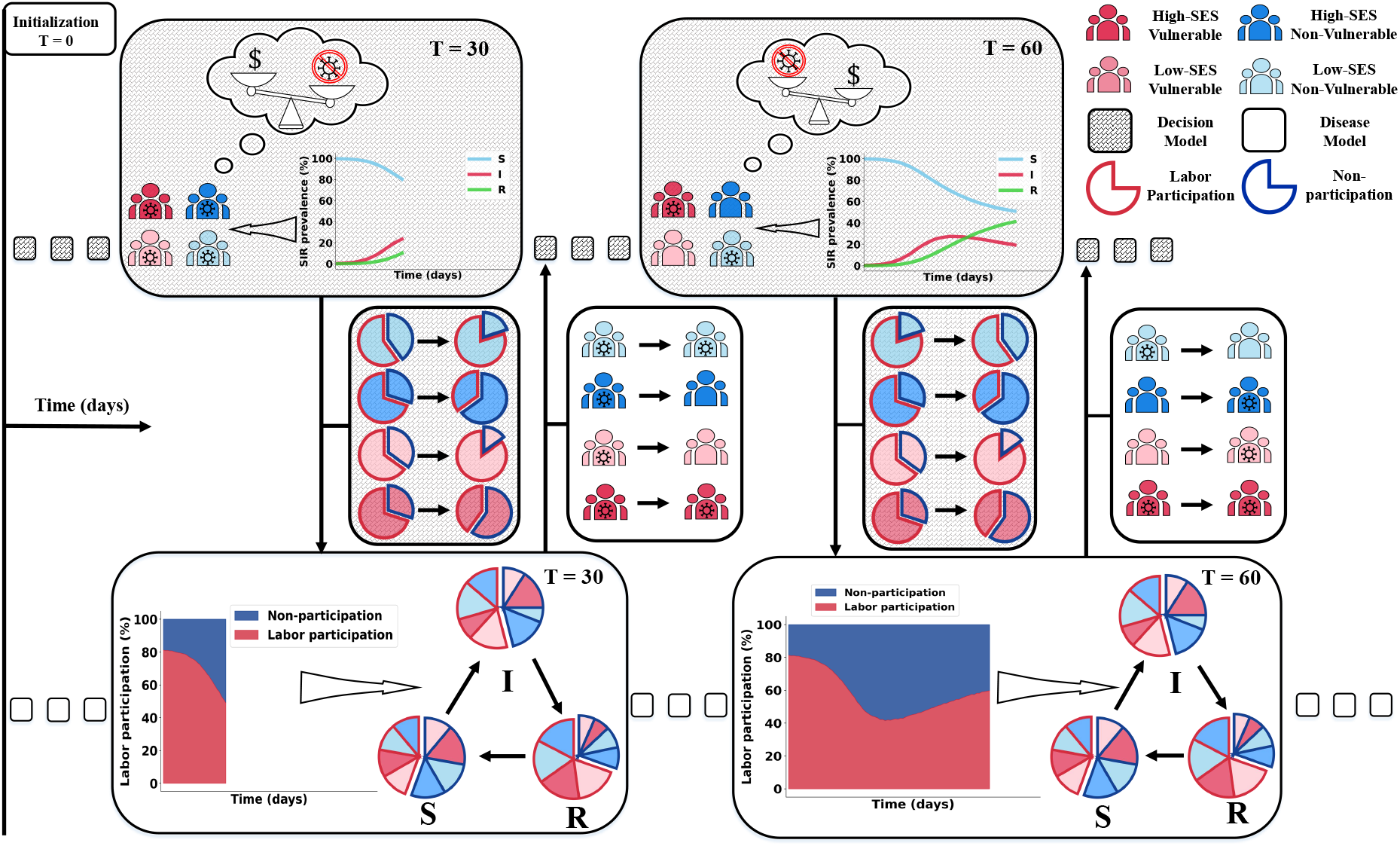
Conceptual overview of the Feedback-Informed Epidemiological Model (FIEM) framework. As an example scenario, we model the decision to work (‘labor participation’) during an outbreak of an infectious disease transmitted between casual contacts. Individuals are stratified by their vulnerability to disease (red vs blue) and socio-economic status (SES) (bright vs pale color). Over time (left to right of figure) they repeatedly weigh the trade-off between income from working and risk of disease. Top row: Decision-making model. Each time period (grey hashed box), individuals make a decision (e.g., to work or abstain from working), based on their own infection status, population-level disease burden, and expected individual utility. Middle row: Decisions influence the distribution of individuals across risk groups (pie charts), which feeds into the epidemiological model (downward arrow). Bottom row: Epidemiological model. Risk groups membership, which depends on individual characteristics and decisions, influences the probability of transitioning between infection states (e.g., S - susceptible/uninfected, I - infected/infectious, R - recovered/immune). The updated individual and population-level disease burden then influences decisions made in future periods (upwards arrow and middle row). The framework is adaptable to other behaviors and population characteristics. See Methods for details.

### Individual-level decision model

Individuals make decisions each period, such as whether to work or not, based on their perception of infection levels and the expectations they form about how their choices influence their future risks of getting infected. Individual decision-making is modeled as a discrete choice to maximize expected lifetime utility, a well-established method in economics (McFadden, 1974; Rust, 1987; Train, 2009) that aligns with other methods of modeling behavior from psychology and sociology (Kubanek, 2017). Individuals make decisions dynamically—their actions are optimal from their individual perspective given how these choices influence the current period’s utility (a function of their infection and non-infection state variables along with their choice) as well as the expected (since future outcomes are probabilistic) present discounted stream of future utility. An optimal decision thus reflects the utility payoffs, information, and beliefs structure of the model, which can be flexibly specified within this modeling framework. Individual and population state variables evolve each period based on the decisions made by individuals in the population.

### Risk-stratified infection model

Each period, individuals in FIEM are classified into a discrete set of risk groups based on their behavioral choices and non-infection state variables (Figure 1). The risk groups are used to construct a stratified compartmental model of infection spread, which tracks at a minimum the proportion of each risk group that is susceptible or infected, but may also track symptom severity, degree of immunity to infection, or diagnostic status, for example. Parameters governing the transitions between disease states can vary by risk group (e.g., contact rates, susceptibility to infection or severe outcomes, duration of infectious period), and individuals may preferentially make contact and thus transmit to others in similar risk groups. The dynamic infection model simulates disease spread and progression to determine the distribution of infection states at the end of each period.

### Disease-decision feedback loop

The core of our model lies in the dynamic feedback loop between individual behavior and the distribution of disease states in the population. Aggregated individual decisions in combination with baseline characteristics then determine the distribution of people across risk groups, which subsequently alters the overall disease dynamics, and thus future optimal individual behavior, going forward. This cyclical process captures the complex interplay: the infection level in the population influences individual-level behavior, and those behavioral responses in turn reshape the trajectory of the disease in the population.

### Decision scenario and parameterization

To demonstrate the capabilities of FIEM, we designed a simple scenario to capture one of the core trade-offs faced during the early stages of the COVID-19 pandemic: the decision to work and earn income or stay home and minimize disease risk (see Methods for details on model structure and parameterization). Disease spread in the population is described by a simple SIRS (susceptible, infectious, recovered, susceptible) model, with an infectious period of *≈* 7 days, average duration of immunity of *≈* 6 months, and a basic reproduction number (*R*_0_) of 2.6. Each period, if an individual chooses to work, they earn income but are more likely to contact infectious individuals, become infected, and incur costs (monetary and otherwise) related to infection. Potential costs of working for susceptibles include missed income or disease symptoms if infected, while infected individuals who work incurs costs such as discomfort of working while sick or stigma against working while infectious. Abstaining from work reduces income, which in turn reduces how much an individual can consume and thus utility from consumption. We include strong health-wealth trade-offs by incorporating two additional margins of individual heterogeneity— socioeconomic status (SES, low or high) and vulnerability to the disease (vulnerable or non-vulnerable). For example, while we assume the average non-vulnerable individual would be willing to pay *∼* $2,000 per day to avoid infection (relative to a mean daily income of $180), a vulnerable individual would be willing to pay triple this amount to avoid infection. If an individual with low-SES chooses not to work they would have to reduce their consumption by 85%, while a high-SES individual would forgo 75% in the same situation. For simplicity, we don’t explicitly model working from home but our parameterization indirectly incorporates its main effect: reducing work contacts is less costly for high-SES individuals. When making decisions, we assume that individuals can accurately assess their own infection status, as well as their short-term risk of infection conditional on both their decision to work and the population-prevalence of infection (which we assume is correct but delayed by a one week lag in case reporting). FIEM can easily accommodate alternative assumptions about the information available to individuals and their understanding or beliefs.

### Dynamic behavior modification alters epidemic trajectory

Awareness of disease transmission in the community triggers individuals to make decisions to reduce the costs of being infected, and our integrated epidemic-behavior model (FIEM) captures this dynamic feedback endogenously (Figure 2). Compared to a traditional “fixed decision” epidemic model where the proportion of the population working is constant (i.e., constant contact patterns) for the duration of the outbreak, under FIEM, workforce participation drops quickly after the outbreak starts, resulting in slower initial epidemic growth and a lower peak (i.e, behavioral feedback naturally “flattens the curve”). Early on, knowingly-infected individuals choosing not to work due to the additional costs of working while infected are the main driver of the reduction in epidemic growth rate, but as infection prevalence increases, susceptible individuals avoid work due to the perceived risk of infection. In both cases, the drop in workforce participation results in fewer contacts between susceptible and infectious individuals and thus fewer new infections. Longer-term, in the absence of additional interventions, the proportion working is predicted to increase again as the peak recedes, but infection to persists leading to a persistently lower working population than before the outbreak. When both the standard and feedback-informed models are parameterized to give the same average labor participation rate (proportion working) over a year-long simulation period, FIEM predicts fewer infections. This simple comparison shows how including endogenous behavior can alter predictions of disease trajectories.

**Figure 2:**
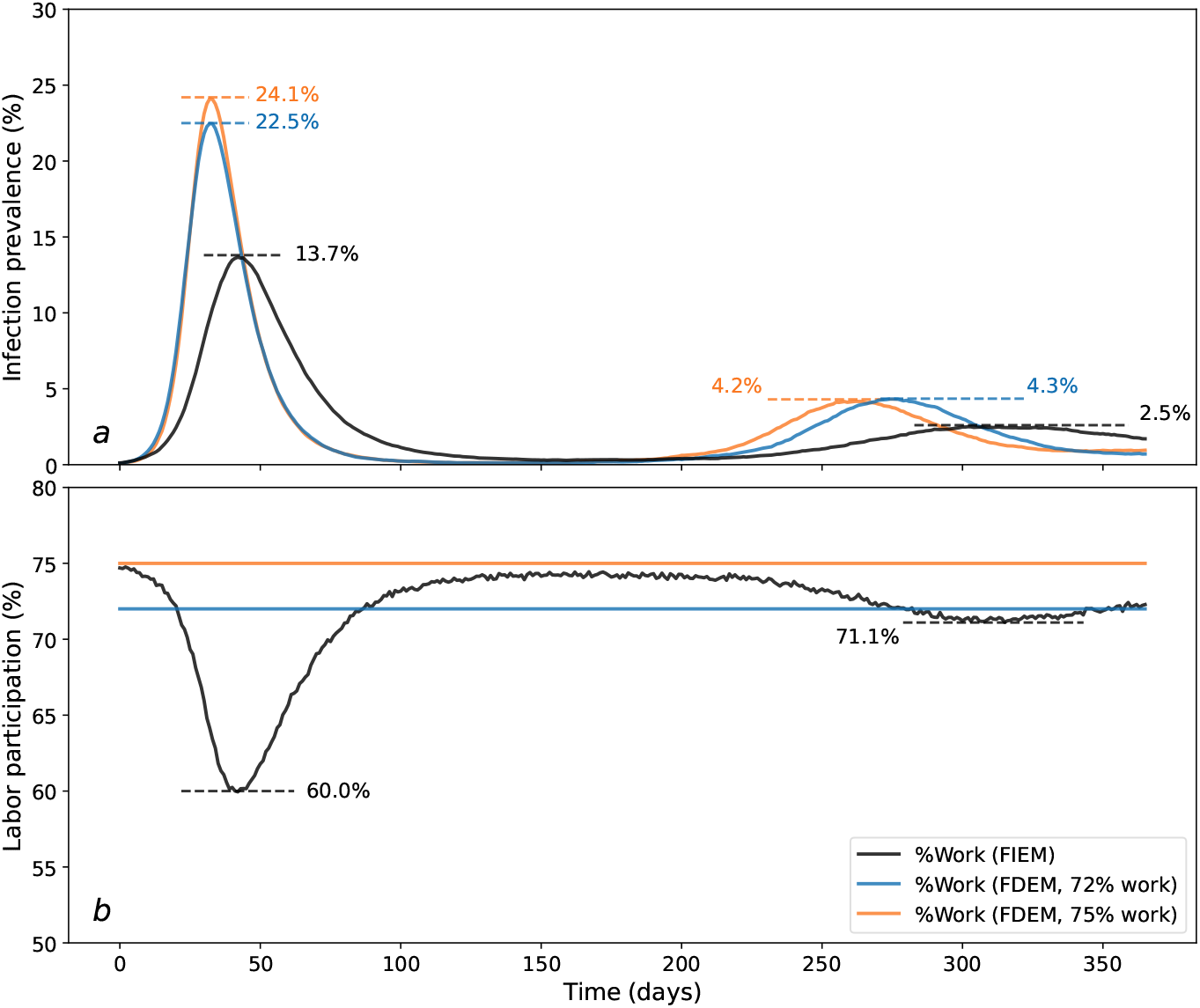
Effect of endogenous behavior change on disease dynamics. The time-course of infection levels and workforce participation for three model scenarios. In the feedback-informed epidemiological model (‘FIEM’, black), individuals dynamically decide to work or abstain from work based on perceived costs and benefits, altering the degree of workplace transmission and thus population-level infection burden (initial working 75%, minimum working 60%, average working 72%, *R*_0_*∼* 2.05, peak prevalence 13.7%). Two alternative fixed decision models are included for comparison: one in which the proportion of the population working each period is held constant at the pre-outbreak level (orange, 75% working, peak prevalence 24.1%, *R*_0_*∼* 2.62), and another where the work level is held constant at the average value observed in the feedback informed model after 1 year (blue; *R*_0_*∼* 2.52, peak prevalence 22.5%, 72%, blue,). *R*_0_ values are estimated by fitting the logarithmic infection curves for the first 20 days. (a) The share of the population that is infected under different models. (b) The share of the population that choose to work each period under different models.

The predicted impact of behavior on disease dynamics depends on the underlying assumptions of the model, in particular, the health and wealth “payoffs” individuals weigh in their decision-making process (Figure 3). Infections that transmit more efficiently cause earlier and higher peaks and trigger earlier and more dramatic reductions in labor participation by susceptible individuals (Figure 3a). If contacts at work are a larger portion of total contacts, meaning the majority of potential exposure to infected individuals occurs at work, a greater proportion of individuals choose not to work. However, within the predefined sensitivity range, the resulting epidemic curve shows no substantial deviation from scenarios where work contacts are less prevalent (Figure 3b).

**Figure 3:**
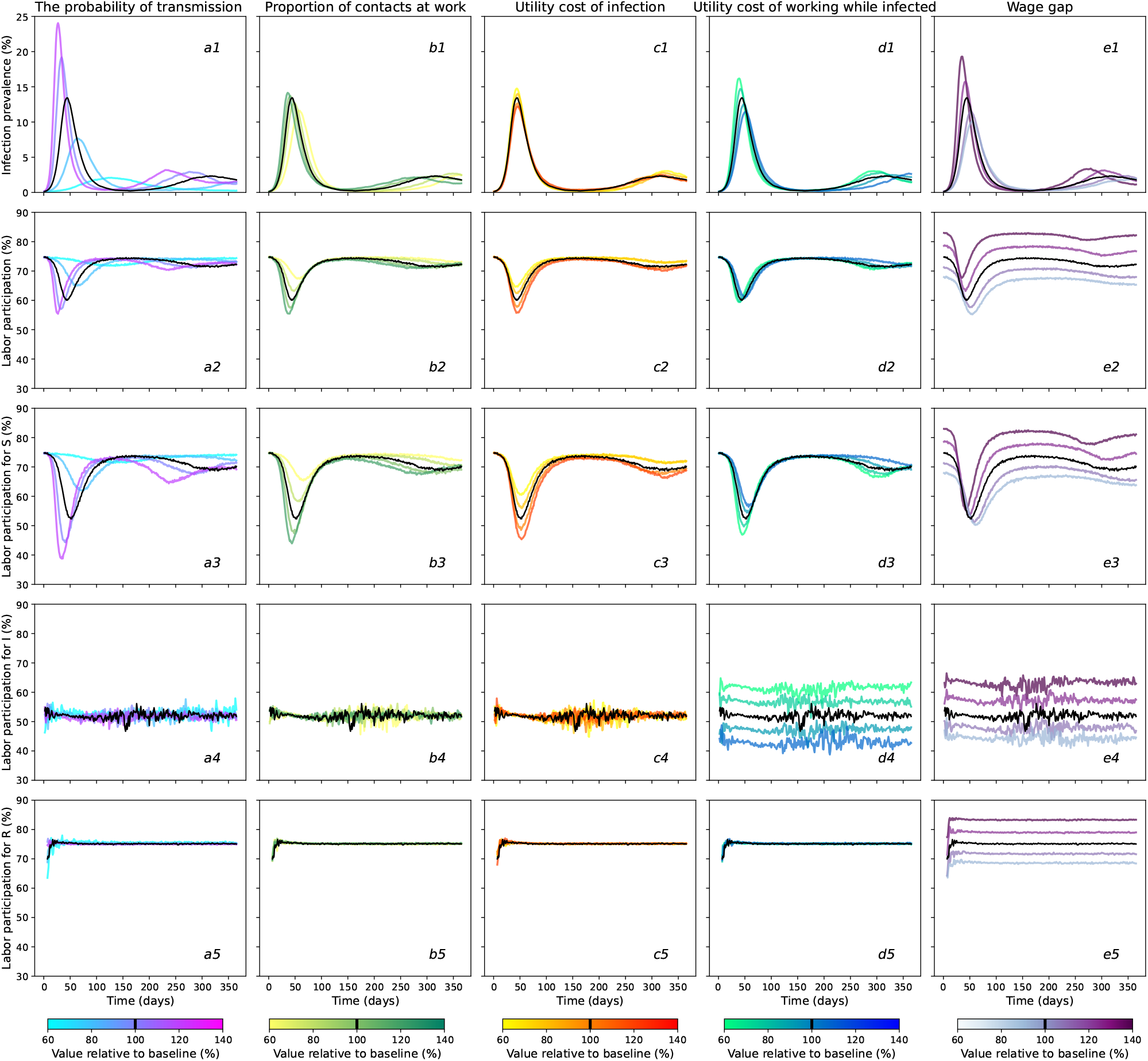
Mechanisms driving individual behavior and disease feedback. The black curves in each panel show our baseline parameter (Figure 2, Tables 2, 3). Row 1) Population infection prevalence. Row 2: Population labor participation. Rows 3–5: Labor participation by susceptible (S), infected (I), and recovered (R) groups. Parameters are varied by column: a) transmission rate, b) proportion of all contacts occurring at work (holding *R*_0_ constant), c) utility cost of infection, d) utility cost of choosing to work while infected, e) the wage gap (the difference in consumption when working vs not working). For all parameters, ranges extend from 40% below to 40% above baseline values

The “utility cost of infection” captures the average cost of being in the infected state, reflecting the experience of typical symptoms of infection as well as rare but costly severe outcomes. As this cost increases, susceptible individuals have stronger incentives to avoid the increased risk of infection at work. Thus, labor participation drops lower once infection becomes common and the epidemic peak is blunted (Figure 3c). In contrast, the “utility cost of working while infected” represents the additional cost of working for infected individuals. As this cost increases, infected individuals are more likely to stay away from work, reducing their contact with others. More infected individuals choosing to stay home leads to a decline in the early epidemic growth rate as well as the peak infection rate (Figure 3d). Importantly, higher values of the “utility cost of working while infected” lead to an *increased* labor supply of susceptible individuals infected individuals optimally choose to abstain from work and the risk of acquiring infection at work thus decreases. Finally, “wage loss” (i.e., the difference between individuals’ income if they do or do not work) further influences work choice decisions. Thus, greater wage losses creates a stronger incentive to work despite illness or risk of infection, since consumption (a component of the utility function) increases with income from wages. As labor participation increases, so does disease transmission, causing larger epidemic peaks (Figure 3e). This pattern is partially driven by the design of this model scenario, which abstracts from financial savings, but we note that this incentive would persist in a model that allowed individuals to reduce the variation in their consumption each period by relying on their savings. Notice that the amount of disease transmission associated with a particular labor participation rate varies with other model parameters, as they influence the incentives for individuals with different health states to work or not. The impact of other parameters— such the time lag in individuals’ information on population-level disease burden and additional utility cost of infection for vulnerable individuals are shown in Supplemental Figures S4 and S5.

**Table 1:** A summary of policy scenarios and resulting health and economic outcomes. Numbers in parentheses represent metrics relative to the no intervention scenario. For each policy, we assume the policy is enacted 20 days after the first infection (when infection prevalence is *∼*2.5%), and continues for 4 months before being relaxed. Total daily cost per capita includes both lost wages due to the disease (compared to a disease-free scenario with 75% labor participation) and the cost of any subsidy payments provided (A detailed breakdown of costs, stratified by wage loss and subsidy payment, is available in Supplementary Table 1). Each subsidy payment amount was benchmarked against the average wage within the population ($180 per day).

| Policy | Description | Policy Scenario | Peak infection prevalence | Average infection prevalence | Average labor participation | Total daily cost per capita |
| --- | --- | --- | --- | --- | --- | --- |
| No intervention | The simulation results were generated from the FIEM without applying any interventions. | No policy applied | 13.7% (1) | 8.2% (1) | 66.7% (1) | \$13.0 (1) |
| Labor restriction | This policy randomly constrains a defined share of the population to remain at home, while the rest are free to choose whether or not to work. | 30% Labor restriction | 6.9% (0.50) | 5.2% (0.63) | 48.9% (0.73) | \$36.3 (2.79) |
| | | 40% Labor restriction | 4.9% (0.36) | 3.9% (0.48) | 43.0% (0.64) | \$44.4 (3.42) |
| | | 50% Labor restriction | 3.6% (0.26) | 3.1% (0.38) | 36.5% (0.55) | \$53.2 (4.10) |
| | | 60% Labor restriction | 2.5% (0.18) | 2.3% (0.28) | 29.9% (0.45) | \$62.1 (4.78) |
| | | 70% Labor restriction | 2.1% (0.15) | 1.8% (0.22) | 23.0% (0.34) | \$72.0 (5.54) |
| Unconditional cash transfer | This policy mirrors the payments the American government provided during the recent COVID-19 pandemic, offering a daily subsidy payment to all individuals regardless of choices. | Payment = 10% average wage | 9.0% (0.66) | 6.5% (0.79) | 59.5% (0.89) | \$37.9 (2.92) |
| | | Payment = 20% average wage | 7.2% (0.53) | 5.6% (0.68) | 55.4% (0.83) | \$60.5 (4.65) |
| | | Payment = 30% average wage | 6.0% (0.44) | 4.8% (0.59) | 53.2% (0.80) | \$81.2 (6.25) |
| | | Payment = 40% average wage | 5.5% (0.40) | 4.5% (0.55) | 51.0% (0.76) | \$101.8 (7.83) |
| | | Payment = 50% average wage | 4.8% (0.35) | 3.9% (0.48) | 50.0% (0.75) | \$121.3 (9.33) |
| Conditional cash transfer | This policy provides a daily subsidy exclusively to individuals who choose not to work, aiming to mitigate the economic consequences of that decision. | Payment = 10% average wage | 7.6% (0.55) | 5.8% (0.71) | 57.5% (0.86) | \$30.0 (2.31) |
| | | Payment = 20% average wage | 5.2% (0.38) | 4.2% (0.51) | 51.8% (0.78) | \$46.3 (3.33) |
| | | Payment = 30% average wage | 4.0% (0.29) | 3.6% (0.44) | 47.3% (0.71) | \$63.1 (4.85) |
| | | Payment = 40% average wage | 3.7% (0.27) | 3.3% (0.40) | 43.4% (0.65) | \$80.6 (6.20) |
| | | Payment = 50% average wage | 2.7% (0.20) | 2.5% (0.30) | 40.8% (0.61) | \$96.6 (7.41) |
| Paid sick leave | This intervention directly targets the health-wealth trade-off infected individuals face by providing financial support, making it less economically harmful to stay home. | Payment = 10% average wage | 10.4% (0.76) | 7.3% (0.89) | 67.3% (1.01) | \$12.2 (0.94) |
| | | Payment = 20% average wage | 8.6% (0.63) | 6.4% (0.78) | 68.1% (1.02) | \$11.8 (0.91) |
| | | Payment = 30% average wage | 7.4% (0.54) | 5.8% (0.71) | 68.8% (1.03) | \$11.5 (0.88) |
| | | Payment = 40% average wage | 6.7% (0.49) | 5.3% (0.65) | 69.3% (1.04) | \$11.4 (0.87) |
| | | Payment = 50% average wage | 5.9% (0.43) | 4.9% (0.60) | 70.0% (1.05) | \$11.3 (0.87) |

**Table 2.** Decision Model Parameters (Θ)

| $\Theta$ | Interpretation | Value | Unit | Ref |
| --- | --- | --- | --- | --- |
| $\theta_x$ | Utility cost of infection. This cost is only paid when an individual's health state is infected. Given $\theta_x = -5$, the average individual in this study would be willing to pay \$5,970 per day to not be in the infected state. See the Parameters section for an estimation of the daily cost of infection in dollars. | -5 | | (Kearsley, 2024) |
| $\theta_v$ | Additional utility cost of infection for vulnerable individuals. This feature is intended to capture the additional incentive vulnerable people have to avoid illness. Relative to the average non-vulnerable person in our simulated population, the average vulnerable individual would be willing to pay triple the amount of non-vulnerable to avoid infection. We intentionally set this value to be large to highlight the role of health-wealth trade-offs in our scenario. | 3 | | |
| $\sigma_h$ | We assume that hassle costs follow a log-normal distribution such that $\log h_{mt} \sim \mathcal{N}(0, \sigma_h)$. This distributional assumption has two implications. First, it imposes that hassle costs are positive. Second, there is a closed form for its expected value, which is computationally convenient. | 0.25 | | |
| $\theta_h$ | Given the $\log h_{mt} \sim \mathcal{N}(0, \sigma_h)$ and $\theta_h = -0.5$, the main hassle cost paid by the average individual is equivalent to \$116 per day, a one standard deviation increase in the hassle cost paid by the average individual in our simulated population is equivalent to \$193, a one standard deviation decrease in the hassle cost paid by the average individual in our simulated population is equivalent to \$73 per day. | -0.5 | | |
| $p_c$ | Our framework assumes that the hassle costs of working are greater for infected individuals. It impacts the labor supply decisions of infected individuals, with an increase in $p_c$ from 0 to 2 reducing the probability of working from 0.75 to 0.55. | 2 | | |
| $w_L$ | Wage for low-SES individuals. The wages for low socioeconomic statuses (SES) were calculated from the 20-40% of family annual income, which is \$35,600 per year, which converts to approximately \$98 per day. | 98 | \$/day | (Board of Governors of the Federal Reserve System, 2024) |
| $w_h$ | Wage for high-SES individuals. The wages for low socioeconomic statuses (SES) were calculated from the 60-80% of family annual income, which is \$95,700 per year, which converts to approximately \$262 per day. | 262 | \$/day | |
| $\bar{c}_L$ | The baseline consumption values are derived based on the amount of family holdings of financial assets for these quantiles. The baseline consumption value implies a savings rate of 15% for low-SES individuals. | 15 | \$/day | |
| $\bar{c}_H$ | The baseline consumption value implies a savings rate of 25% for high-SES individuals. | 66 | \$/day | |
| $\kappa$ | Discount factor. It captures the value of future flow utility payoffs today. A value of 0.96 implies individuals tend to prefer payoffs today but are still willing to wait for future period payoffs. A discount factor of 0.96 implies an annual discount rate of 4%. | 0.96 | /day | (Frederick et al., 2002) |
| $\nu$ | Euler's constant | 0.57721 | | |
| $l$ | Information lag for each individual aims to capture the delay between onset and case reporting. | 7 | days | (Abbott et al., 2020) |

**Table 3.** Infection Model Parameters (Ψ)

| $\Psi$ | Interpretation | Value | Unit | Ref |
| --- | --- | --- | --- | --- |
| $\beta$ | The probability of transmission per infectious contact per time. This parameter is calibrated to give $R_0 = 2.62$ using the model with fixed decisions with 75% individuals working. | 0.025 | /day<br>/individual | (Dhungel et al., 2022) |
| $\gamma$ | Rate of recovery from infectiousness $g$, based on estimates of the serial interval and the duration of shedding of infectious virus centered at approximately 1 week. There is substantial variation in estimates across studies, individuals, and metrics, and infectious period is expected to depend on control measures that lead to isolation of infected individuals. | 0.14 | /day | (Byrne et al., 2020; Ali et al., 2020; Hakki et al., 2022; Lavezzo et al., 2020) |
| $\alpha$ | Rate of waning of protective immunity after natural infection. Two systematic reviews measured the risk reduction for re-infection in individuals with documented prior infection, and suggest protection decays to 50% at 6 months, corresponding to an effective waning rate of $\ln(2)/180$ days. | 0.004 | /day | (Stein et al., 2023; Bobrovitz et al., 2023) |
| $C_0$ | Baseline total number of contacts of each individual. | 4 | | (Mistry et al., 2021; Mossong et al., 2008; Prem et al., 2017) |
| $\sigma_1$ | Increased propensity to come into contact with individuals of the same vulnerability and socioeconomic status (“preferential mixing”). | 1.5 | | (McPherson et al., 2001; Newman, 2002; Smith et al., 2014; Hilman et al., 2022) |
| $\sigma_{\text{work}}$ | Increased propensity to come into contact with other working individuals when deciding to work. | 4 | | (Mistry et al., 2021; Mossong et al., 2008; Prem et al., 2017) |
| $\sigma_{\text{SES}}$ | Increased propensity of contact between individuals of low SES. Low-SES people tend to work in industries with more in-person contacts and experience more household crowding. Value abstracted from studies to represent an intermediate degree of assortativity. | 1.5 | | (Papageorge et al., 2021; Dingel and Neiman, 2020; Nande et al., 2021; Jay et al., 2020; Kamis et al., 2021; Emeruwa et al., 2020) |
| $\mathbb{C}$ | The contact matrix $C$ is a function of $C_0, \sigma_1, \sigma_{\text{work}}, \sigma_{\text{SES}}$. | Eq. (14) | | |

*Notes:* This table lists the model parameters, their interpretation, and calibrated value that we use in our scenarios. Unless explicitly noted, model parameters are fixed at these values.

### Consequences of heterogeneous health-wealth trade-offs

To demonstrate our framework is capable of accounting for the inherent heterogeneity of real-world populations, we next examine how variation among individuals in vulnerability to disease and socioeconomic status impact behavior, shape trade-offs, and subsequently influence the epidemic trajectory. High-SES individuals make more money if working and have a lower opportunity cost of not working, which we attribute to omitted factors, such as savings or having jobs that allow work-from-home arrangements. Low-SES individuals have more contacts at work and preferentially contact other low-SES individuals. Vulnerable individuals face higher utility costs from infection (i.e., have a higher likelihood of progression to more severe infection), but have no difference in per-exposure susceptibility to acquiring disease. We evaluated the infection trajectory predicted by our feedback-informed model for a baseline population with an even distribution of individuals across four risk groups (i.e., non-vulnerable/high-SES, vulnerable/high-SES, non-vulnerable/low SES, vulnerable/low-SES) (Figure 4, see Figure S6 for alternative distributions yielding similar results).

**Figure 4:**
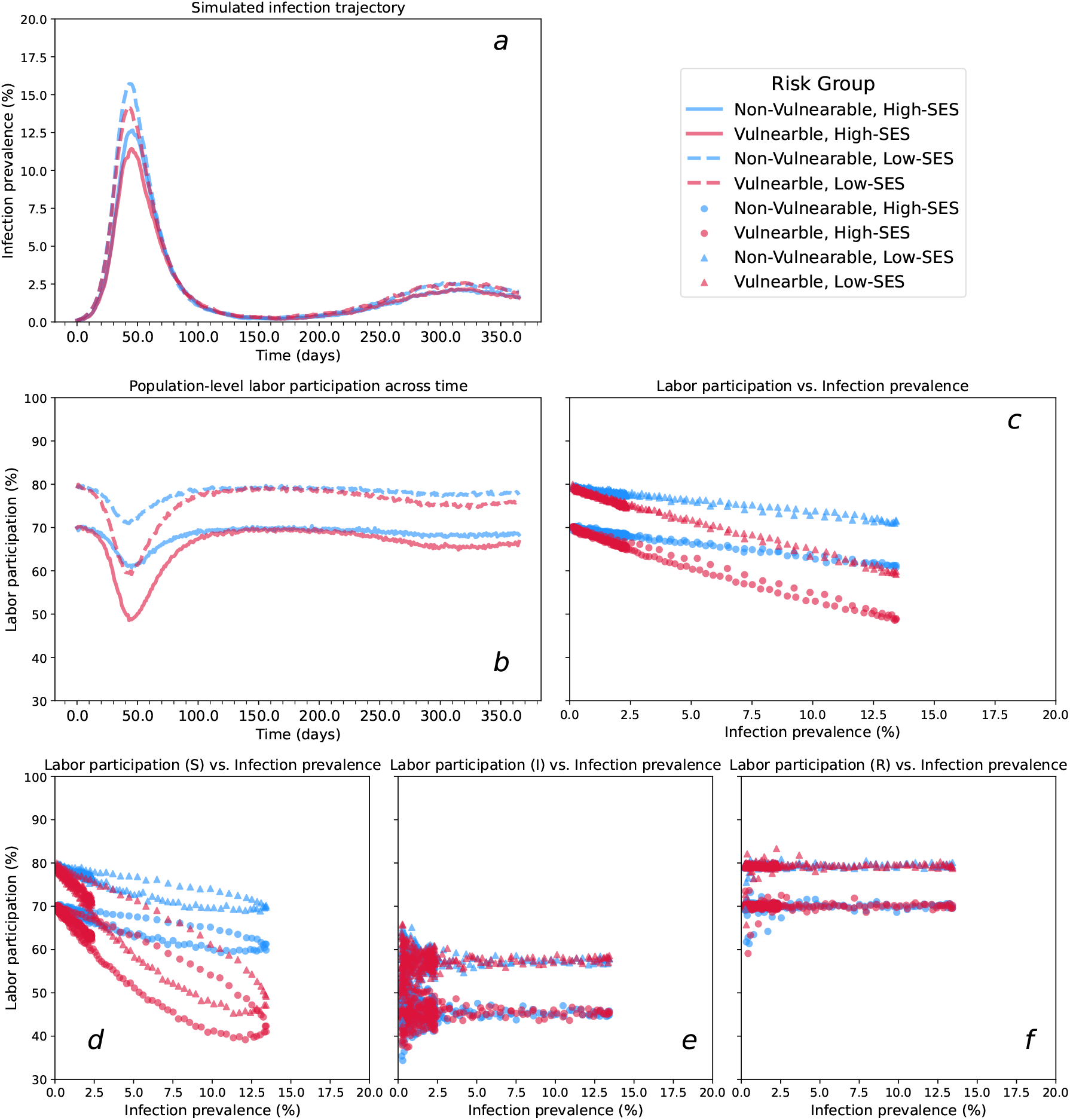
Disease-decision dynamics across heterogeneous risk groups. The dynamics of infection levels and the decision to work in a population stratified by vulnerability to severe disease if infected (vulnerable - red, non vulnerable - blue) and socioeconomic status (high SES - solid line or circle, low SES - dashed line or triangles). a) Infection prevalence over time; b) Labor participation (fraction working) over time; c) Labor participation vs infection prevalence for all infection statuses (d–f) Labor participation vs infection prevalence for susceptible (S), infected (I), and recovered (R) individuals. In this scenario, the four risk groups are of equal size.

We start by analyzing individual incentives to preserve their economic well-being. Low-SES individuals face a stark and disproportionate trade-off between economic needs and health preservation. Low-SES individuals choose to work during the early outbreak stage despite infection risk, driven by their urgent need to meet necessities. As a consequence, low-SES individuals experience higher early exponential growth rates and epidemic peaks (Figure 4, dashed curves and triangles). Conversely, high-SES individuals exhibit more cautious behavior, with more individuals abstaining from work for a given infection level, reflecting their financial ability to prioritize health over wealth (Figure 4, solid curves and circles). These results underscore the need to consider socioeconomic inequalities when designing public health policies.

Next we analyze individuals’ incentives to protect their health. Vulnerable populations exhibit stronger self-protective behavior due to the higher risks associated with infection. This feature creates an incentive for these individuals to not work, forgoing some consumption, during the period of highest disease burden (Figure 4, red curves and shapes). Since we assume individuals have perfect information about their current health state, only the susceptible group responds to infection risks (Figure 4d–f). This assumption can be relaxed to capture settings where infected individuals may be unaware of their status and either i) abstain from work believing it could prevent infection, and ii) continue to work without experiencing any of the utility costs of working while infected.

Our results highlight how key differences in health-wealth trade-offs experienced by different risk groups influence the joint trajectory of infection and behavior, as well as distributional consequences of the epidemic.

### Evaluating the impact of policy interventions

The goal of our framework is to provide a tool for analyzing disease control policies that incorporate endogenous behavior changes to improve prediction of infection burden, understand the distributional consequences of policies, calculate welfare, and identify optimal policies based on the specific needs and values of decision makers. To illustrate this potential, we encoded and compared four different policy interventions within our model (Table 1, Methods): *labor restriction*, in which a portion of the population is constrained to abstain from work; *unconditional cash transfer*, in which a daily subsidy payment is provided to all individuals; *conditional cash transfer*, in which a daily subsidy payment is provided exclusively to individuals who choose not to work, and; *paid sick leave*, in which infected individuals who choose to abstain from work receive baseline wages and additional subsidy payments. We assume perfect compliance with policy recommendations and perfect knowledge of infection status.

We compare peak infection and labor participation outcomes under varying degrees of intervention for each policy (Figure 5). Labor restrictions have the largest marginal impact in reducing total and peak infections. The highest level of restriction we simulate (70%) lowers peak prevalence from 13.7% in the no-intervention scenario to 2.1% (Table 1, Figure 5a), but carries large economic burdens; translating to a $59 average loss of income per day per capita relative to a scenario where the disease outbreak occurs with no infection control policies in place and endogenous behavior change. Unconditional and conditional cash transfer policies also demonstrate considerable reductions in peak infection rates; at the highest payment levels (50% of the average wage), peak prevalence is reduced to 4.8% for unconditional and 2.7% for conditional transfers (Figure 5b–c). These policies result in higher labor participation than labor restrictions, with unconditional transfers maintaining higher participation than conditional transfers. However, cash transfers have higher direct costs to the government than labor restriction. Paid sick leave tends to have less impact on reducing peak and average infections than cash transfers, but it increases the average share of the population choosing to work (*≈* 70%, vs 50–60% for the unconditional transfer and 40–58% for the conditional transfer).

**Figure 5:**
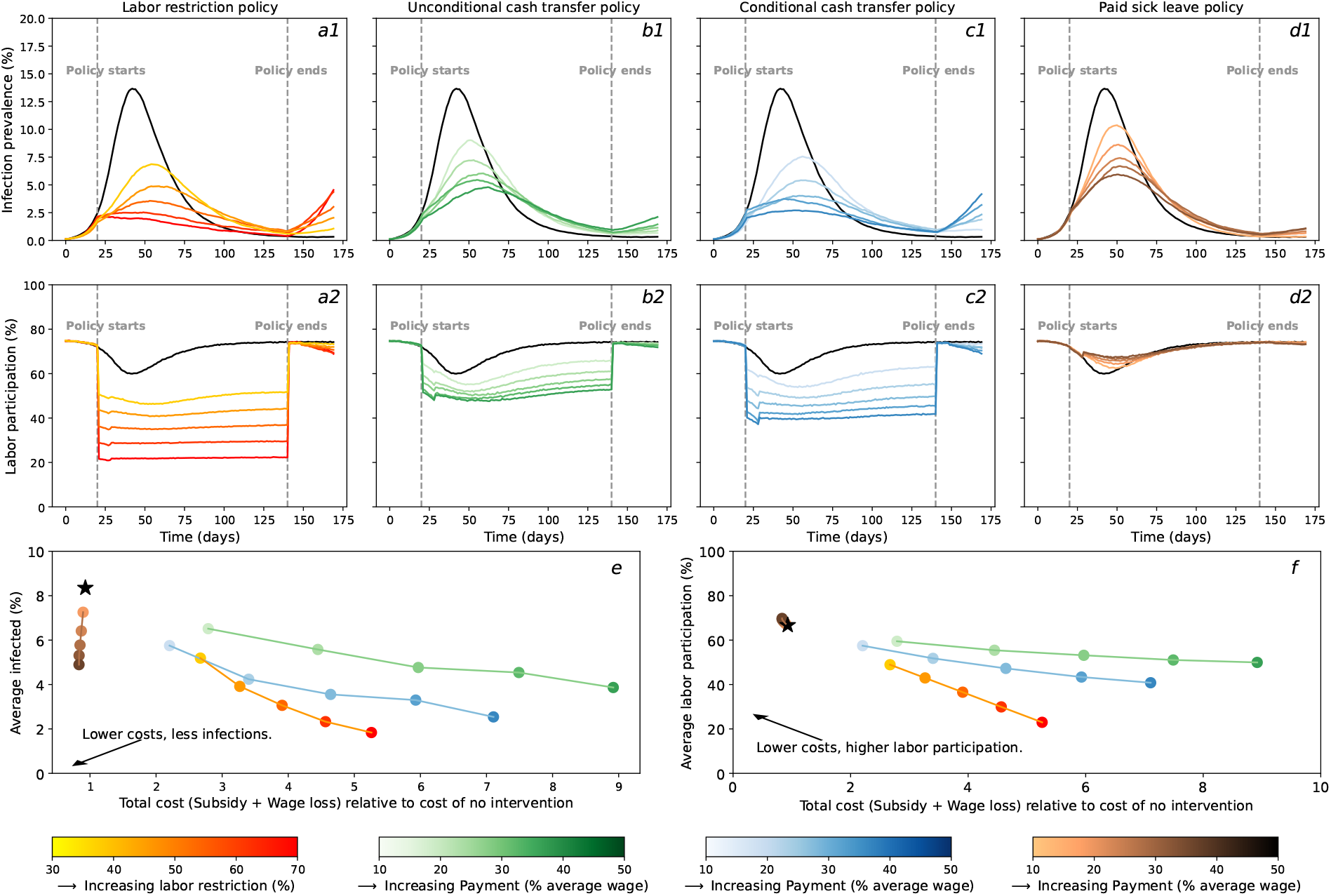
Effects of policy interventions on population health outcomes and labor participation. a-d) Population infection prevalence and share of the population choosing to work over time, for different policies. The black curve is the baseline model with no intervention—no labor restriction and no additional payment. (a) Labor restriction policy (red to yellow) that limits how much of the population is able to choose to work. (b) Unconditional cash transfer (green) that is delivered to all individuals each period. (c) Conditional cash transfer (blue) that is delivered to individuals who choose not to work each period. (d) Paid sick leave policy (brown), which allows infected individuals to earn their full wage if they choose to not work while infected. The simulations start with no policy intervention in place. The intervention begins on day 20 and remains in place for 4 months (until day 140). e) Average share of the population that is infected while policy is in place versus cost of policy. e) Average share of the population chosing to work while policy is in place, vs cost of policy. Policy costs includes the sum of government spending to fund transfers and lost wages to individuals relative to their baseline wage earnings predicted by the model when there is no disease present.

We also evaluate the cost-effectiveness of each policy. Costs are defined as the net of subsidy payments and wage losses due to reduced labor participation, and expressed both as a dollar value and percentage relative to the no intervention scenario (Table 1, Figures S6 and S7). We did not include other potential costs associated with these policies, such as the costs of administration, diagnostic tests, or enforcing restrictions. We evaluate effectiveness in terms of the peak infection prevalence, but other epidemiological metrics could also be used. For labor restriction and cash transfers, stronger versions of the policies which incur higher costs are associated with lower peak infection levels. Labor restrictions achieve equivalent peak infection reductions for lower costs than other policies (Figure 5e–f). For example, under the example parameters used for this simulation, a labor restriction policy costing around $40 per person per day reduced peak infections to a third of the no intervention scenario, whereas achieving similar reductions costs close to $120 with an unconditional cash transfer. However, the paid sick leave policy deviates from this pattern, and uniquely achieves reduced infection rates and lower total costs as subsidy payments increase. For example, providing 50% of the average wage for a paid sick leave policy reduces the total daily cost per capita to $19 (0.76 of the cost with no intervention) and the average infection rate to 5.9% (0.43 of the peak size of no intervention). Paid sick leave accomplishes this infection reduction while also *increasing* the average amount of labor supply in the population, thereby reducing wage loss costs. By giving infected individuals a strong incentive to not work, the risk of infection for a susceptible person declines and allows them to endogenously decide to work, an example of a positive externality of the policy. In reality, the cost-effectiveness of paid sick leave is complicated by the issue of accurate detection of infectious individuals and malingering. However, our framework’s ability to model individual decision-making allows us to capture the core effects of this policy and could be expanded to include more details, providing valuable insights for policymakers who must consider both the intended and unintended consequences of their interventions.

We also use our framework to evaluate which policy designs are optimal for achieving pre-specified objectives (Figure 6). To do so, we construct a social welfare function, which specifies how to weigh the cost of the policy versus the benefit of fewer total person-days of infection and whether to impose a budget constraint for the policy’s costs. These components may vary across scenarios or across policymakers. Once the social welfare function is specified, we can solve for the policy stringency or payment level that maximizes this function subject to its constraints. We demonstrate how to perform this type of analysis with the conditional cash transfer policy. We use a weight that reflects a willingness to pay $1 per capita per day to reduce the total number of people infected per day by 2.71, which is based on the value per statistical case of COVID-19 used by the US Department of Health and Human Services (Kearsley, 2024). Given these conditions, we find the optimal policy is a $65 payment per individual per day. Defining the social welfare function for optimal policy is a complex decision, but FIEM can flexibly capture different weights or budget constraints policymakers must contend with when analyzing and designing policy.

**Figure 6:**
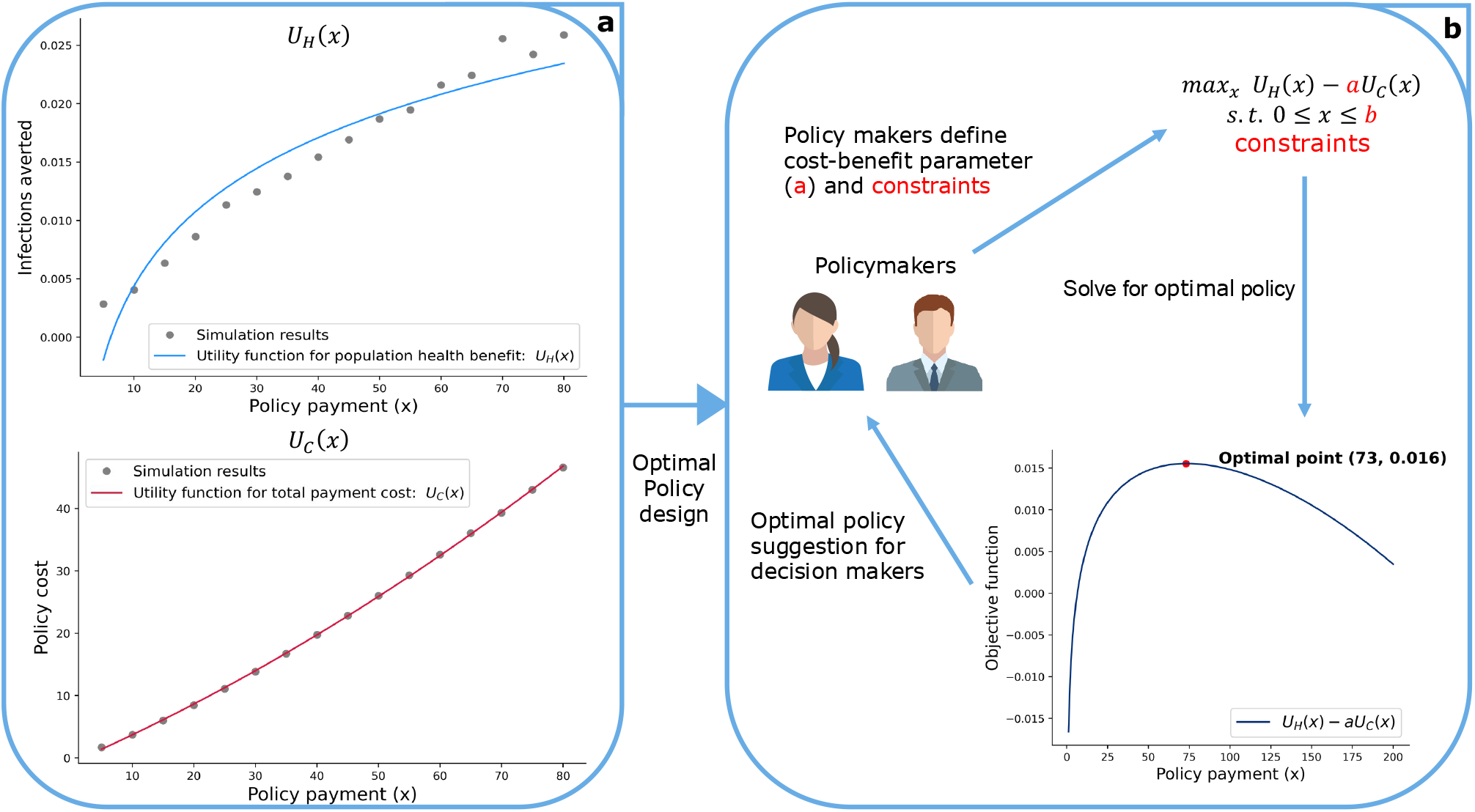
Decision-maker informed optimal policy design. Schematic diagram outlining how policymakers can use the feedback-informed epidemiological model to identify an optimal policy design. We use the unconditional cash transfer policy as an example, assessing its impact over a 4-month implementation period. a) The simulated health benefits *U*_*H*_ (*x*) and policy costs *U*_*C*_ (*x*) as functions of daily subsidy payment, fitted to generate continuous curves. Health benefits *U*_*H*_ (*x*) is defined as the reduction in the average fraction of days an individual spends infected during the evaluation period. This is calculated as 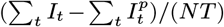, where *I*_*t*_ represents the number of infected individuals on the day *t* with no intervention, 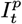 *N* represents the number of infected individuals on the day *t* with implemented policy, *N* is the population size, and *T* is the duration of the evaluation period. The policy cost *U*_*C*_ (*x*) is defined as the payment cost per capita per day over the evaluation period, given by (∑_*t*_ *C*_*t*_)*/*(*NT*), where *C*_*t*_ denotes the policy cost on day *t*. Increasing policy payment increases both health outcomes and the associated cost of the policy, creating a trade-offs between the two. b) Policymakers decide how to numerically weigh the relative benefits and costs of each policy, and specify any monetary or political economy constraints. This allows for the definition of a single objective function *U* (*x*) = *U*_*H*_ (*x*) *− aU*_*c*_(*x*) that can be maximized to determine optimal payment amount, subject to the assumed parameters and defined constraints. In this example policy optimization, we estimated the daily cost of infection per capita to be $5,912 (see Method parameters section for a detailed calculation) so that a unit increase in *U*_*C*_ (*x*) ($1) would be equivalent to 0.00017 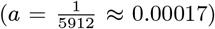 of a unit in *U*_*H*_ (*x*). Increasing *a* places greater emphasis on minimizing policy cost, while decreasing it prioritizes reducing infections averted. Note that the estimated values above are dependent on the length of the evaluation period. In this example, the optimal policy would be identified as a $73 per person per day cash transfer, which would be expected to avert approximately 22 days of infection per 1,000 individuals in the population.

### Distributional consequences of policies in heterogeneous populations

To evaluate the differential impact of policy interventions in subgroups experiencing different health-wealth trade-offs, we assess impacts of each policy by socioeconomic status and vulnerability to disease (Figure 7). Consistent with population level outcomes, we found that all policies effectively reduced infection levels in all groups, but failed to eliminate disparities in infection burden by SES status, although differences between the groups were slightly reduced for more stringent policies. However, we observed heterogeneous behavioral responses. Subsidy-based interventions disproportionately influence the behavior of low-SES groups, moving from the no-intervention scenario where they are more likely to maintain high labor participation despite infection risk to creating opportunities for them to acknowledge their higher infection risk at work and abstain. For example, conditional cash transfers cause a sharper decline in work participation for the non-vulnerable, low-SES group (from 59% to 35%) compared to the high-SES group (60% to 48%). Interestingly, the vulnerable, high-SES group exhibits modest reductions in choosing to work. This pattern is partially driven by the reduced labor supply of low-SES individuals, which makes the probability of infection lower for a high-SES individual if they opted to work. As the paid sick leave subsidy increased, labor supply is maintained despite reductions in infection, and this is largely driven by vulnerable populations, consistent with a reduction in their health-wealth trade-offs.

**Figure 7:**
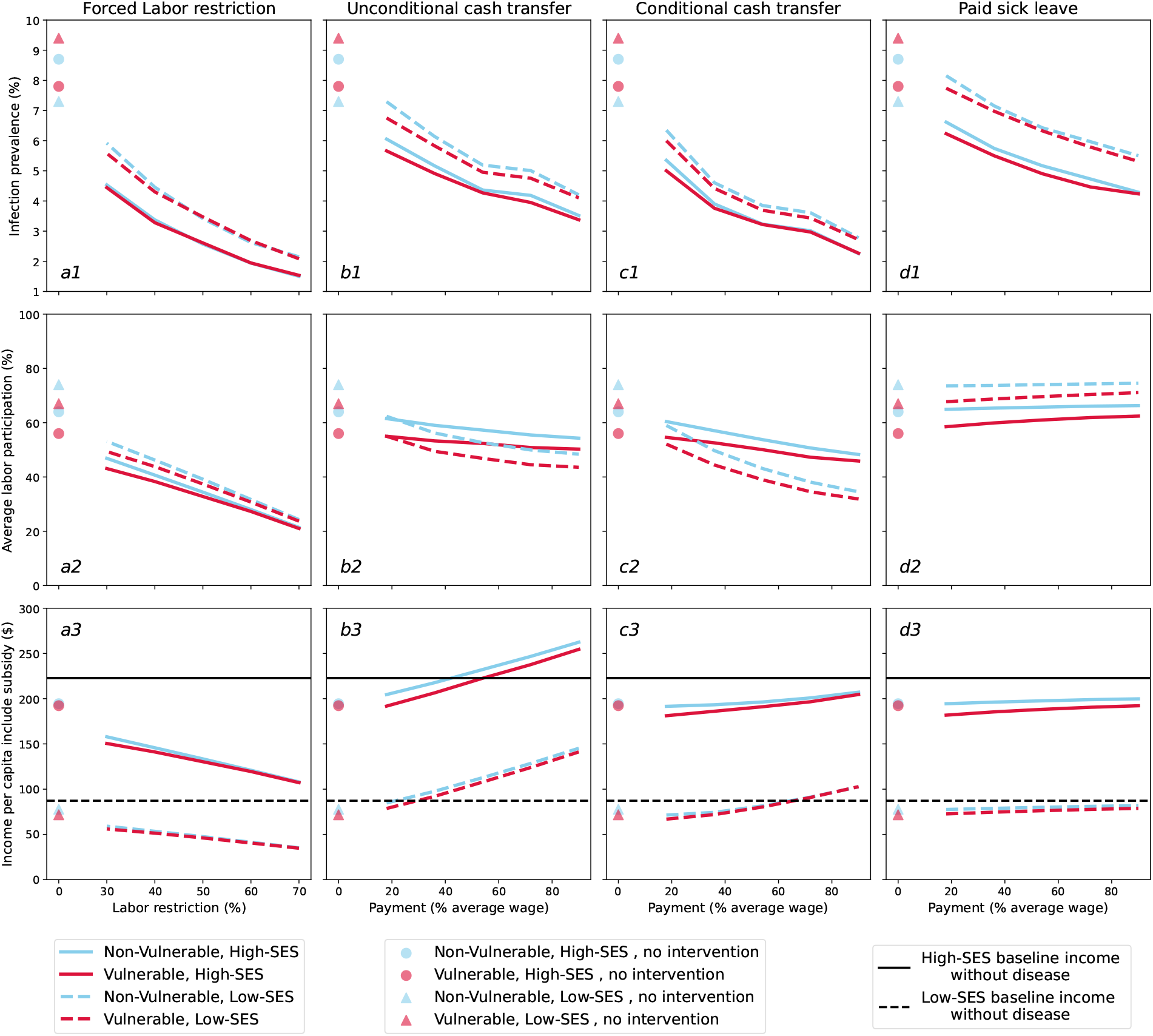
Heterogeneity of policy impacts by risk groups. Row 1: average population infection rate. Row 2) average labor participation Row 3: income per capita. a) Labor restriction policy limiting how much of the population is able to choose to work. b) Unconditional cash transfer delivered to all individuals each period. c) Conditional cash transfer delivered to individuals that choose not to work each period. D) Paid sick leave allowing infected individuals to earn their full wage if they choose not to work while infected. Heterogeneity in outcomes across risk group are denoted by different curves: non-vulnerable (blue), vulnerable (red), low-SES (dashed lines and triangles), and high-SES (solid lines and circles). The black line gives baseline income without disease.

We also compare total income under each policy intervention relative to baseline income by population subgroups. Each policy generates notable differences. For example, low-SES individuals benefit substantially from the unconditional cash transfer policy relative to high-SES people. This pattern is driven by the size of the transfer relative to their labor earnings. Conversely, conditional cash transfer offers slightly higher benefits to high-SES individuals when payments are low, as they already have less incentive to work. Paid sick leave provides marginal but similar benefit to income for all groups, by increasing the share of the susceptible population of each group that opts to work. In contrast, labor restrictions significantly disrupt labor supply and negatively impact the income of individuals in all risk groups, with larger drops among low-SES individuals. This highlights how policies that appear effective by population-average metrics, like overall infection prevalence reduced per dollar of total cost, may exacerbate existing disparities and thus be considered suboptimal from an equity standpoint.

## Discussion

During a public health crisis, policy makers must balance population health and economic well-being. This task is further complicated by subgroups of the population facing economic precarity or health vulnerability (or both), so any policy will have unequal distributional consequences. The goal of useful models is to predict the impacts of policy so that at minimum, we avoid inefficient policies, e.g., those with higher economic costs for no additional health benefits, or those with higher health burden but no economic benefits. In this paper we present a flexible modeling framework that captures the feedback between individual decision-making and infectious disease spread. We integrate a mechanistic model of disease dynamics (consistent with established best-practices in infectious disease epidemiology) with a formal model of individual decision-making based on forward-looking utility maximization (commonly used in economics). Our “feedback-informed epidemiological model” (FIEM) can flexibly encode the processes by which an individual’s perceived risk of infection, among other factors, influences their behavior, which in turn impacts future disease propagation. Our long-term goal is to use FIEM—following retrospective validation with diverse data sources—to provide multi-objective forecasts and scenario projections to public health policymakers who must consider both health and economic consequences of infectious disease outbreaks and their control.

To illustrate the capabilities of FIEM, we designed a simplified scenario inspired by the early stages of the COVID-19 pandemic. Individuals decide whether to work or not based on the trade-off between health and economic well-being—abstaining from work lowers income but also reduces the risk of infection. Results of these simulations show that our model can endogenously propagate the implications of this decision process, which leads to slower epidemic growth and reduced peak disease burden but fails to achieve control. By systematically varying the parameters governing the disease-behavior feedback, we identify multiple channels through which this feedback operates, and gain an understanding of their relative importance (Figure 3). In the real world, individuals experience different trade-offs between preserving health and maintaining economic well-being, and we expanded our scenario to stratify individuals by both their vulnerability to severe disease and their socioeconomic status (Figure 4). Our model predicts behavioral responses consistent with these trade-offs—individuals with lower SES that are less vulnerable to the disease are most strongly motivated by the need to preserve their economic well-being, leading them to maintain the highest rates of continued in-person work during an outbreak and experience the highest peak infection levels. Similarly, high-SES individuals that are vulnerable to the disease have a strong incentive to preserve their health and thus are more likely to reduce working in-person. This example scenario ignores many features of COVID-19 driven decisions, for example the high proportion of asymptomatic infections, limited testing, misinformation, the role of accumulated savings, the inability of many workers to leave and re-enter the labor force at will, and the purely pro-social motivations of some individuals for engaging in costly disease-avoidance behavior. However, even in a simplified model, our analyses underscore how individualized health-wealth trade-offs and dynamic decision-making contribute to behavior change at the individual level, which carries direct implications for aggregate disease spread.

Continuing with our COVID-19-inspired scenario, we use our model to evaluate the effects of four policies aimed at reducing disease spread: labor restrictions, unconditional cash transfers, conditional cash transfers, and paid sick leave. These policies were evaluated against a policy with no intervention but allowed for endogenous decision-making that informed the path of the pandemic. Each policy reduced peak and total infections, albeit through different mechanisms and at different costs to individuals and the government (Figure 5a–d). For instance, labor restrictions significantly lowered infection prevalence but required individuals to forgo substantial consumption, while cash transfer policies achieved comparable reductions in infection but necessitated considerable government expenditure to counteract individuals’ incentives to work (Figure 5f). FIEM also captures externalities induced by government policies. For example, paid sick leave inadvertently increased the number of people choosing to work during an outbreak by effectively isolating infected individuals. Finally, we demonstrate how this framework can identify optimal policy designs that balance the benefits of reduced infections with the associated policy costs (Figure 6), and decompose the heterogeneous impacts these policies have across different sub-populations (Figure 7). We believe this framework has the potential to advance policy-relevant disease modeling in multiple ways, including i) simultaneously outputting epidemiological, microeconomic, and macroeconomic metrics, ii) incorporating the impact of risk-avoidance behaviors that occur independently of mandated behavior change, and iii) centering considerations of equity in policy projections, by producing sub-group specific impacts.

Despite the many simplifying assumptions in the model scenario presented here, the structure of FIEM allows it to be extended in many directions. Our behavioral model makes two key assumptions: First, individuals engage in dynamic utility optimization with a low discount rate, thereby excluding other behavioral patterns such as impatience, hyperbolic discounting, and alternatives to dynamic optimization. Second, we assume individuals possess rational expectations (i.e., can accurately assess their current health status and future risk the pandemic will evolves). However, in reality more complex decision models may be at play along with uncertainty over information about the outbreak. These assumptions can be tested with data on human behavior, beliefs, or information transitions, and FIEM can be readily adapted to incorporate these features as warranted. An additional assumption is the exclusion of strategic interactions among individuals. While it is possible to relax this feature, doing so may lead to multiple equilibria in the behavioral model, introducing methodological and computational complexities. Our model currently only considers individual decisions, whereas businesses and other institutions (e.g., schools) also engage in risk-avoidance decision making in response to disease. More critically, our current framework is not designed to describe macroeconomic processes that may also feed back with individual decision making during public health crises, such as changes in labor demand, economic growth or recession, inflationary processes, interest rate changes, among others. However, FIEM can incorporate macroeconmoic models and produce integrated forecasts.

The epidemic model considered here was deliberately simplified to highlight concordance with classic compartmental models, limit the number of parameters, and facilitate interpretation of results. However, FIEM can easily include more complex disease dynamics and health outcomes. For example, we could extend the model to track individuals’ knowledge of their infection status (via testing, symptoms, etc.); imperfect reporting, access, and interpretation of data on population-level disease burden; decisions that impact not only contact probabilities but susceptibility to infection, duration of infectiousness, or propensity for severe disease; prosocial behavior in which individuals incur a cost to avoid transmitting disease to others even in the absence of individual risk; and capacity constraints to healthcare resources. Our framework currently classifies individuals into a defined set of strata depending on the combination of their infection state, risk factors, and health decisions, but could readily be extended to an individual-based model, albeit at substantially increased computational cost.

Our model captures the components essential for credible prediction of disease spread and endogenous behavioral responses in a way that is often omitted by other attempts to integrate these two features (e.g., disease spread models with substantial heterogeneity but no explicit individual-level optimization informing behavior, or economic models of human behavior with non-standard epidemiological processes Fenichel, 2013; Eichenbaum et al., 2021; Acemoglu et al., 2021; Ash et al., 2022; Haw et al., 2022; Pangallo et al., 2023). However, past work has included other important details that were omitted here but could be integrated into future work. For example, Brotherhood et al. (2023) present a rich behavioral model, which captures detailed decisions about time use (i.e., leisure in and outside of home, work in person, and teleworking), the production and consumption of different types of goods (i.e., leisure goods outside of home and consumption within home), and model parameters that are at least partially fitted to match real world data. Another example is Ash et al. (2022) that presents a coupled epi-economics model that formally captures how financial constraints enter the individual’s decision problem. Finally, the framework proposed by Pangallo et al. (2023) effectively replicates the economic and epidemiological factors of a specific geography. These features can improve the quality of the forecasts and predictions a model produces. While FIEM does not currently incorporate these features, future work can extend the framework to capture these valuable model components.

The decision to abstain from work to avoid infection was particularly salient during the early phase of COVID-19, when rapid at-home tests, medical-grade face-masks, and vaccines were unavailable. Our model could be extended to consider the decisions that became more important as these interventions were introduced. These decision models will likely be more complex, as individuals incur “costs” beyond lost income that may be harder to quantify—such as stigma, social isolation, inconvenience, discomfort, or irrational fears. For other sorts of infectious diseases, different decision paradigms arise—to adhere to long-term, nausea-inducing drugs to prevent eventual disease progression or transmission; to lose a potential romantic partner by disclosing an STI, and so on. Our framework allows for many extensions in these directions, and we anticipate that the limitation to including them will not be the ability to encode a reasonable model within the FIEM structure, but to identify data appropriate for estimating model parameters. Here we have merely “calibrated” our model—choosing a single reasonable parameter set that roughly recreates a small set of aggregate epidemiologic or economic metrics. Future work will present methods for integrating diverse datasets for formal inference of FIEM parameters. We hope that case studies using this framework will provide the motivation for behavioral and microeconomic data collection as a core component of pandemic preparedness activities, so that future disease-behavior models can produce more informed policy recommendations and include uncertainty intervals in all projections.

## Methods

### Model Overview

#### General framework

In this paper we present an integrated dynamic framework that captures human behavior and disease spread that we refer to as the Feedback-Informed Epidemiological Model (FIEM). This complex exercise involves fully integrating methods from two distinct fields in a reasonable, credible, and tractable way that does not unduly limit either dimension. From epidemiology, we use a risk-stratified dynamic compartmental model of disease spread (Anderson and May, 1991; Keeling and Rohani, 2008). From economics, we draw on methods rooted in dynamic utility maximization and discrete choice modeling (McFadden, 1974; Rust, 1987; Train, 2009). These components are integrated by allowing behavior at the individual-level to influence aggregate-level outcomes such as disease spread. An advantage of FIEM is that it is flexible and can readily incorporate extensions in all three of these dimensions (i.e., the model of behavior, the epidemiological disease model, and aggregate outcomes). In this section, we provide a basic description of the key model components. It is written for an audience familiar with mathematical concepts used in economics and epidemiology so that members of each discipline understand our decisions and crucially how we integrated the tools from each discipline.

We are concerned with a population of individuals exposed to an infectious disease. At each time point (period), each individual is characterized by a set of “state variables” that describe their infection status, economic well-being, and other factors that may influence health or wealth going forward. At fixed times, individuals make a behavioral decision based on calculating how each possible decision will impact their overall well-being (“utility”, a function of state variables and their decision) in the current period, and predicting how it will impact their state variables—and thus utility—in future periods. These decisions may involve trade-offs—between different aspects of utility like health vs wealth, or between current and future well-being—and optimal choices may vary between individuals. For example, a person with pre-existing health conditions might see infection as more costly than a person without pre-existing health conditions and optimally choose to engage in less risky behavior to avoid getting infected. Similarly, a person with low-socioeconomic status might see a greater downside to not working, since the amount they can consume (i.e., income not saved) if they do not work is very low. The forward-looking aspect of an individual’s decision is captured by how they perceive the likelihood of infection. If they observe low infection levels, they may conclude that engaging in riskier behaviors is worthwhile. This is the channel through which disease dynamics influence choices (in the current period).

After individuals have made a behavior decision, they are aggregated into risk groups (or more generally, assigned an individualized risk-level), defined by an individual’s state variables and the decisions they made for the current period. Within the model, a period is a point in time when individuals make decisions about their behavior and it can be flexibly adjusted based on the context (daily, weekly, etc.). Risk group membership determines how individuals experience different states of infection and how likely they are to transition between different states (e.g., the number of social contacts and thus risk of acquiring infection, duration of infectiousness, severity of symptoms, likelihood of knowing their infection status, or efficacy of therapy). Given the distribution of risk groups and the distribution of infection states in each, the epidemiological model predicts how the pathogen will spread among the population in a given time window, and thus determines how an individual’s infection status changes from the current period to the next one. Guided by the epidemiological model, infection transmits probabilistically from infectious to susceptible individuals, and infected individuals progress through different stages of infection. This process, along with transitions for other state variables in the model, determines what state variables the individual will observe in the next period when the decision process begins again.

FIEM integrates behavior and disease spread models in two ways. First, individuals react to their own as well as population-wide infection levels—described by the epidemiological model—when making choices about what actions to take in the current period. Second, while individuals make individually-optimal decisions, these choices are aggregated across the population and used as inputs that affect how the infection evolves from one period to the next. As a result, FIEM is a complete model that captures the feedback between behavior and disease spread. These are the two features that are critical for evaluating the effects and heterogeneous burdens of counterfactual policies for infection control.

#### Key Components

FIEM has several key model components, which are briefly summarized below and expanded upon below in the **Model Details** section delow.

The first component is the flow utility function *u*(*z, d, ϵ*; Θ), which measures the overall well-being (utility) of an individual in the current period. Utility (*u*(*·*)) is a function of observable-state variables summarized by *z*, the decision(s) the individual makes denoted by *d*, state variables unobserved to the modeler but known to the individual *ϵ* and a vector of utility model parameters Θ. Each period, individuals make decisions to maximize the present discounted value of their lifetime flow utilities based on how they expect state variables to evolve in the future, given their actions in the current period.

The next component of FIEM is the set of rules governing the transitions of state variables over time, and the expectations individuals have about them, described by transition matrix **P**(*z*^*′*^, *ϵ*^*′*^| *z, d, ϵ*; Ψ) with transition model parameters Ψ and primes denoting the state variables for the next time step. We break this component down into transitions involving non-infection state variables (**A**) vs changes in infection status (**Q**). Non-infection state variables could change due to processes such as aging, relocation, acquisition of medical conditions that impact disease risk, job loss or promotion, or emergence of new pathogen variants that have differential risk profiles. Infection status transitions include progression from being in an uninfected to infected state, or infected to recovered state. Transitions could be deterministic or stochastic, and may or may not depend on the actions of others individuals in the population. Individuals may have perfect or imperfect information about the nature of these transitions depending on the application and available data.

The innovative component of FIEM is our framework governing transitions of infection status state variables, which endogenously incorporates outcomes from the modeled decision process. The dynamics of infection are described by a risk stratified compartmental model of disease spread. Individuals acquire and transmit infection, and progress between stages of disease, according to established epidemiological principles and pathogen-specific parameter values. The incidence of new infections depends jointly on the prevalence of susceptible and infectious individuals, and on their likelihood of contact, so individual transition probabilities are always tied to the infection status of others in the population. The risk group an individual belongs to can directly influence their probability of transiting between disease states, by way of risk-group specific parameters (e.g., vulnerability effects risk of developing severe disease; decision to work impacts number of contacts). In addition, heterogeneous contact patterns across risk groups—non random mixing—can allow for concentration of infection in certain risk groups and amplify differences in infection dynamics between groups (e.g., low SES individuals have increased risk of acquiring infection due to higher number of contacts, and the fact that those contacts are more likely to be infected). The risk group assignments of individuals, and hence the distribution of risk groups in the population, are determined by the decisions individuals make as governed by the behavioral model as well as other state variables. This creates a dynamic feedback between disease spread and behavior.

Together, the utility function and the expectations about state variable transitions between time periods form the final key object of FIEM, the value function *V* (*z, ϵ*; Θ, Ψ). This function captures the dynamic decision problem by describing the total expected utility—the sum of the current period utility plus (discounted) expected values in future periods given possible state transitions—under the assumption that individuals will make optimal decisions in each period. The value function can be solved using Bellman’s principle of optimality (Bellman, 1957; Dixit, 1990) and associated numerical methods, to extract expressions for choice probabilities conditional on current state variables. These rules allow FIEM to predict behavior that arises under different policies and, when combined with data on decision making, can be used to facilitate the estimation of the utility function’s parameters, which are econometically identified under modest assumptions about the distribution of unobserved state variables, the discount rate, and individual preferences (Hotz and Miller, 1993; Magnac and Thesmar, 2002).

#### Scenario and Parameters

To demonstrate the capabilities of FIEM in this paper, we designed a simple scenario based on the acute phase of the COVID-19 pandemic. Our goal is to capture and evaluate two forms of vulnerability that impact decision making and disease spread: health and economic. Our approach features calibrated model parameters, informed (but not directly inferred) from the literature. We demonstrate our calibrations are reasonable as the model reproduces predictions consistent with basic theoretical premises of epidemiology and economics (e.g., if more people engage in risky behavior, more infection will happen; people work if doing so means they earn more money; etc.). In future work we can expand the model to include additional details, formally infer parameters from data, and propagate parameter estimate inference to the model’s predictions, for example to test whether predicted differences between policies are statistically distinguishable from zero. Disease spread is described by a simple SIRS model (susceptible, infectious, recovered, susceptible) model, in which individuals who are “susceptible” to disease may transition to become infected based on exposure to another infected individual, and those who are infected are “infectious” and capable of transmitting infection for some time, before progression to a “recovered” state where they are no longer infectious and have developed some immunity to re-infection, which may eventually wane, leading them to return to the susceptible state. We used an average infectious period of 7 days, an average duration of fully-protective immunity of 6 months, and a basic reproduction number (*R*_0_) of 2.6 (average number of secondary infections produced by each infected individual before recovering) (Table 3).

We focused on a single decision that represents a common choice individuals in many settings faced early on in the pandemic. Each period, individuals make a decision to work in person or to not work. If choosing to work, individuals earn income but are more likely to incur costs (monetary and non-monetary) related to infection. Susceptible individuals who work increase their number of contacts with potentially infectious individuals, and thus increase their risk of infection. Getting infected carries utility costs associated with disease symptoms or missed economic opportunities. Working incurs additional costs for infected individuals, which may capture the discomfort of working while sick or the stigma against working while infectious. The value of *R*_0_ we report corresponds to the value when a fixed 75% workforce participation rate, reflecting the population-level average proportion working under the selected wage and baseline consumption without the influence of disease, and assuming no behavior changes in response to the disease.

We allow for two forms of individual heterogeneity in addition to infection state: an individual’s socioeconomic status (SES, high vs low), and their vulnerability to the disease (vulnerable vs non-vulnerable). Labor income and consumption values are parameterized such that not working presents a greater relative trade-off for low-SES individuals than high-SES ones based on data from the Survey of Consumer Finances (Board of Governors of the Federal Reserve Board, 2023). Low-SES individuals are assumed to have more contacts even after conditioning on their decision, and are more likely to contact other low-SES individuals. Vulnerable individuals face higher utility costs of infection compared to non-vulnerable ones (e.g., could experience more severe or longer duration symptoms), but have no difference in per-exposure susceptibility to acquiring disease. We don’t explicitly model working from home, but its impact is approximately captured by our parameterization, which allows high SES individuals to have fewer contacts at work and a lesser decline in consumption if abstaining from in person work. We assume that individuals have perfect information about their own current infection state, but their information about the distribution of infection states within population has a seven day lag. Given this information, we assume individuals can accurately predict how their work decision today will influence their probability of transitioning between infection states in the next period.

#### Assumptions and Extensions

There are several assumptions within FIEM. Unless noted otherwise, these assumptions are common to the FIEM framework and not the specific scenarios in our analysis.

First, we assume individuals engage in dynamic utility maximization with exponential discounting (i.e., we rule out hyperbolic discounting or impatience in decision making). This assumption is testable with data on human behavior or discounting and can be easily adapted to account for these other types of behaviors if warranted. FIEM can also alter the information available to individuals about state variables and how they may evolve given their actions. Our scenario assumes individuals possess perfect information (i.e., correct beliefs) about their risk group and infection state, and that individuals know their probability of transitioning infection states from one period to the next based on the transmission probabilities from seven days earlier. This assumption is intended to capture lags reporting information about disease transmission in the population. Moreover, we require that individuals assume that these transmission rates will hold for all future periods when making their current period choices. In practice this means individuals do not learn to anticipate these updated transmission probabilities at the start of each period. Given data on beliefs or information transition, this assumption can be altered to capture information asymmetries and perception biases, which had a known impact on the response to COVID-19 (Druică et al., 2020; Fragkaki et al., 2021; McColl et al., 2021). Our general framework and scenario abstract from peer effects (i.e., how choices of other individuals influence your decisions) but this could be captured as part of the flow utility function. The general framework and our scenario exercise also rule out the strategic interaction among individuals when making their behavior decisions but do capture how the aggregate levels of infection within a period can influence individual behavior. Relaxing this assumption is possible but can lead to multiple equilibria in the behavior model, which carries methodological and computational challenges. The scenario also assumes that risk-group membership directly informs the probability of contact between individuals. This linkage is key to our framework, though the specifics of these relationships can be modified according to the states modeled and the questions at hand.

Within the FIEM framework, certain assumptions are made regarding the epidemiological dynamics. Primarily, disease transmission is modeled in our scenario using a compartmental structure, meaning individuals are classified into a finite number of discrete infection states, abstracting from the reality in which pathogen load, complex immune responses, and diverse symptoms may continuously evolve in unique ways in each individual. Our particular COVID-19-inspired scenario assumes a novel disease entering a population with no prior immunity, where infection implies infectiousness, and a temporary period of perfect immune protection exists but wanes post-infection (SIRS model), leading to damped oscillations even in the absence of any behavioral feedback. However, our general framework could accommodate any sort of infectious disease. The current FIEM framework assumes that the disease transmission model can be described with a transmission matrix which at each time describes the probability an individual in state *X* will move to state *Y* by the end of the time period (i.e., a Markov process). However, the model could be extended to allow state transitions to also depend on past states (i.e., infection history). The version of the model used in our scenario assumes that the parameters of the disease transmission model are constant over time, but this could be relaxed to allow for dynamic parameter adjustment, for example due to other interventions altering transmission rates (e.g., mask use) or duration of immunity (e.g., pathogen variant).

In our scenario we permit two margins of individual heterogeneity to inform behavior decisions and disease spread. While this makes our scenario simple, FIEM itself is general and can be extended to capture additional features. On the epidemiological side for example, incorporating additional margins like age and co-morbidities can better inform disease spread and health outcomes. Additionally, a SIR model without stratification by age is too rudimentary to adequately capture the dynamics of COVID-19, and does not capture additional complexities like human mobility, emergent pathogen variants, and vaccination which may be relevant to the disease dynamics (Leung et al., 2021; Dyson et al., 2021; Alleman et al., 2023). On the behavior side, the scenario assumes away many factors that are relevant for determining behavior in response to infection (e.g., gender, race, education, risk preferences, political or religious beliefs), omits savings, financial behavior, and interactions between individual behavior and the macroeconomy. Despite these assumptions, we see our scenario exercise as a proof of concept that illustrates how much can be accomplished with a parsimonious model that has these components implemented in a credible manner. The clear next step for future work is to expand out this framework to capture a richer setting, which FIEM is future proofed to do in many directions.

### Model Details

#### Variables

Individuals are indexed by *m*, and time is measured in discrete increments with each period denoted by *t*.The total size of the population at time *t* is denoted by *N*_*t*_ such that *m ∈ {* 1, …, *N*_*t*_ *}*. This notation is flexible and can allow the total size of the population to change over time. In each period *t*, an individual makes a decision *d*_*mt*_ *∈ 𝒟* where *𝒟* denotes the set of possible choices. We use 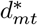 to denote the optimal decision an individual can make within a period. A decision is optimal if it is associated with the highest benefits—where these benefits are measured by a value function—to an individual at that time.

Different individuals may have different optimal decisions within a time period and the optimal decision for individuals need not correspond to the optimal decision for the population. Certain behavioral choices can be associated with costs which are denoted by *h*_*mt*_. These costs are specific to the behavior choice and distinct from other types of costs that may influence other factors such as income lost or infection risk. For example, models of labor supply typically include a term for the disutility of work, which captures the non-monetary costs associated with going to work (e.g., arranging childcare, time spent commuting, interacting with colleagues). The exact interpretation or use of these costs can depend on the application at hand.

State variables refer to the factors that influence the well-being or utility for an individual and may vary over time. In general, there are three types of observable state variables in FIEM. First are individual non-infection factors (e.g., age, socioeconomic status, vulnerability to a disease), denoted by the vector *k*_*mt*_ ∈ *K*, where *K* is the set of all possible vectors of non-infection state variables. The second state variable tracks an individual’s infection state and is denoted by *x*_*mt*_ ∈ *𝒳* and depends on the epidemiological parameterization that is applied. The third type of state variables within FIEM are population-level state variables that are denoted by the vector *E*_*t*_. At a minimum, this vector aggregates the total number of people within each of the infection states in the model but could be extended to track other population-level factors that are relevant for an individual’s well-being (e.g., aggregate economic output in the economy such as wages and good production, vaccine availability, hospital capacity, government policies). We use the vector *ż*_*mt*_ = *{k*_*mt*_, *x*_*mt*_, *E*_*t*_*}* to summarize these observable state variables associated with an individual at a given point in time.

We additionally assume there may be state variables that are known to individuals and impact their decisions, but unobserved to the modeler. We denote these by *ϵ*_*mdt*_; these may be choice specific—hence the *d* index. Including these unobserved variables allows us to recreate the ubiquitous finding that individuals with the same apparent state/information do not always make identical decisions.

FIEM also has outcomes and payoff variables that are related to the decisions made by individuals in the model. Individuals are mapped to risk groups based on their non-infection state variables *k*_*mt*_ and behavioral decisions *d*_*mt*_. An individual’s risk group assignment at time *t* is denoted by *g*_*mt*_ *∈ 𝒢*, where *𝒢* is the set of all possible risk groups. The population-level state variables *E*_*t*_ can be further disaggregated by risk group, denoted as *E*_*gt*_. Individuals’ decisions and state variables influence their utility (see next section) through intermediate variables that are realized or calculated using the particular model structure and parameters relevant to the problem. For example, in our example we track earnings *w*_*mt*_, as well as the subset of earnings spent that influence utility: “consumption”, *c*_*mt*_. Individuals also realize hassle costs each period *h*_*mt*_ that influence their behavior in that period.

#### Decision model

The utility an individual in the model derives at time *t*, which is a measure of “well-being”, is captured by the flow utility function *u*(*z*_*mt*_, *d*_*mt*_, *ϵ*_*mdt*_; Θ), which depends on their observable-state variables *z*, the decision the individual makes denoted by *d*, state variables unobserved to the modeler but available to the individual *ϵ*, and a vector of utility parameters Θ. The exact parameterization of the flow utility function carries implications for how agents will behave by altering their trade-offs and perceptions of risk. These features can vary based on the context or scenario.

In this framework, each individual makes the decision each time period that gives the maximum expected value of their utility over time, conditional on their current state variables. The goal is to determine this optimal decision sequence—or more formally, to solve for the optimal decision in each time period given state variables—under our model for decision-dependent payoffs and state variable transitions between time periods. This dynamic optimization problem is described by the value function,

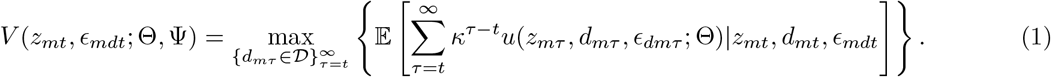

The value function *V* (*z*_*mt*_, *ϵ*_*mdt*_; Θ, Ψ) represents the maximum flow utility payoffs on an infinite horizon ranging from *t→ ∞*, made by choosing the optimal set of future decisions 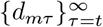. Discounting (*κ <* 1) is used to reflect individuals’ preferences for more immediate payoffs (“present-discounted”). The expectations operator E is used to capture the expected utility under stochastic dynamics governing how the observed and unobserved state variables in future periods (*z*_*mτ*_, *ϵ*_*mdτ*_) will evolve conditional on the current period’s state variables (*z*_*mt*_, *ϵ*_*mdt*_) and behavior decisions *d*_*mt*_. More generally, the expectation could also be over alternative beliefs and individual holds about the likelihood of these transitions. Indicated by the conditional term “| *z*_*mt*_”, this equation includes the constraint that the state variables evolve from one period to another according to the transition rules of the model.

The value function encodes the solution to a dynamic discrete time optimization problem, but solving for an infinite set of future decisions is intractable. To overcome this issue, we use Bellman’s principle of optimality to reformulate the problem as a recursive “Bellman equation” (Bellman, 1957; Dixit, 1990). In simple terms, Bellman’s principle of optimality states that if you’re trying to find the optimal path to a goal, any point along the way should get you closer to your end goal most effectively. This allows us to express the solution of the value function in terms of two consecutive periods—rather than tackling the full infinite sequence at once—thereby significantly simplifying the problem,

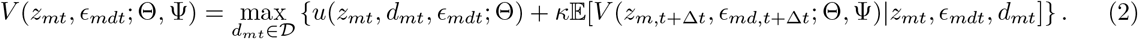

The expected value of the value function over the horizon *t* + Δ*t→ ∞* can be rewritten as the weighted average over all future states (observed and unobserved), where each future state’s contribution is weighted by its probability of occurrence upon transitioning from *t → t* + Δ*t*, as encoded in the transition matrix **P**,

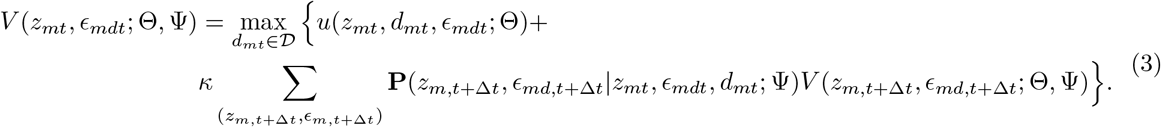

The value **P**(*z*_*m,t*+Δ*t*_, *ϵ*_*m,t*+Δ*t*_ | *z*_*mt*_, *ϵ*_*mt*_, *d*_*mt*_; Ψ) describes the probability that an individual ends up in the state *z*_*m,t*+Δ*t*_, *ϵ*_*m,t*+Δ*t*_ at time *t* + Δ*t* conditional on being in state *z*_*mt*_, *ϵ*_*mt*_ at time *t* and making the decision *d*_*mt*_. In this work, **P** encodes the infection model that describes how individual infection state variables (*x*_*mt*_) evolve based on the actions of individuals within the model. The transition matrix can include components describing transitions for other observed state variables, such as age, and can encode assumptions about how unobserved state variables influence transitions. The transition matrix used in the decision model and epidemiological model can differ, allowing FIEM to capture a situation where individuals base their decisions of faulty perceptions of their transition probabilities.

To make the simulation of the decision model tractable, we must avoid integrating over the unknown states. Hence, we make a series of convenient assumptions about the unobserved state variables *ϵ*— established in the economics literature—to permit calculation of the value function and the optimal decision (Hotz and Miller, 1993; Rust, 1987, 1994; Magnac and Thesmar, 2002; Arcidiacono and Ellickson, 2011):

- Additive separability in the flow utility function, i.e. *u*(*z, ϵ, d*; Θ) = *u*(*z, d*; Θ) + *ϵ*(*d*).
- Conditionally independent in the transition model, meaning that: 1) Conditional on the current observed state and decision choice, the future state does not depend on the current unobserved variables, and 2) conditional on the future state, the future unobserved variables do not depend on the current unobserved variables. This yields: **P**(*z*_*m,t*+Δ*t*_, *ϵ*_*m,t*+Δ*t*_|*z*_*mt*_, *ϵ*_*mt*_, *d*_*mt*_; Ψ) = **p**(*ϵ*_*m,t*+Δ*t*_|*z*_*m,t*+Δ*t*_; Ψ) **p**(*z*_*m,t*+Δ*t*_|*z*_*mt*_, *d*_*mt*_; Ψ).
- Independent and identically distributed across individuals, periods, and choices, so that *p*(*ϵ*_*mt*_| *z*_*mt*_; Ψ) = *p*(*ϵ*; Ψ), and following a type-I extreme value distribution (standard Gumbel distribution). Note that this assumption implies conditional independence.
- Individuals do not forecast how the arguments of *P* will evolve in future periods when making decisions in the current period. Rather, they assume that transitions implied by the current period state variables and transition matrix will persist in future periods and they do not anticipate that this transition matrix will change based on their actions.

While methods exist to relax the assumptions regarding unobserved state variables, they significantly complicate calculations. Under these assumptions, we can define a decision specific “net-of-errors” expected value function 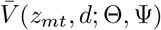 that excludes the decision-specific randomness from the original value function,

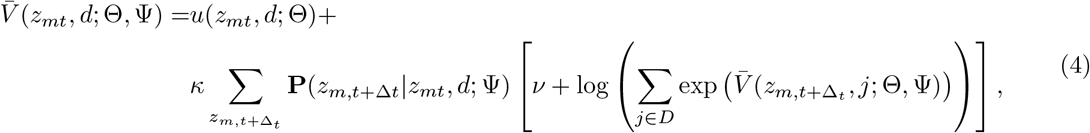

Here *ν* is Euler’s constant, which is the expected utility resulting from the Gumbel distributed unobserved states (*≈* 0.57, distinct from Euler’s number *e≈* 2.72). The assumption that the unobserved states are i.d.d. samples of a type I extreme value distribution conveniently yields an expression for the probability that a particular choice *d* is optimal—and thus chosen—action for individual *m* conditional on the observed state variables *z*_*mt*_,

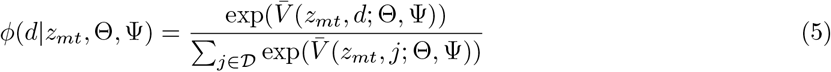

Conceptually, the probability a choice is made is proportional to its relative value; decisions associated with higher values are more likely to be chosen.

If the net-of-errors value function is known, then the conditional choice probabilities of Equation (5) can be used to calculate individual’s decisions at any time, and thus to simulate the model (for example to make predictions under different counterfactual scenarios). They can also be used in a likelihood framework to estimate the parameters of the model. However, Equation (4) does not immediately give the net-of-errors value function, since it is a recursive functional equation. Additional steps are required to find a numerical value function that satisfies this equation, and the method we use to do this is described below in Algorithm 1.

#### Non-infection state variable transitions

Non-infection state variables may evolve from one period to the next. The exact nature of these transitions is application specific. For example, if the scenario calls from tracking an individual’s age across periods, the transition for this state variable may be deterministic. These transitions are allowed to depend on the actions of individuals within the model. For example, in a model that allows for financial savings, the amount of savings in the next period may depend on whether an individual earned labor income in the current period.

To formally capture these transitions, recall **A** denotes the transition matrix for the non-infection state variables *k*, and is a constituent of **P**. The probability that an individual in state *k*_*mt*_ at time *t* transitions to state *k*_*m,t*+Δ*t*_ by time *t* + Δ*t* is given by **A**(*k*_*m,t*+Δ*t*_|*k*_*mt*_, *d*_*mt*_; Ψ), referred to as **A**_*mt*_ for brevity.

#### Infection model

The dynamic epidemic model consists of a discrete set of disease states, *𝒳* and a transition matrix **Q**, whose elements specify the probability that an individual in infection state *x*_*mt*_ at time *t* transitions to infection state *x*_*m,t*+Δ*t*_ by time *t* + Δ*t*, formally denoted **Q**(*x*_*m,t*+Δ*t*_ |*x*_*mt*_, *g*_*mt*_, *E*_*t*_; Ψ), or abbreviated as **Q**_*mt*_. The variable *g*_*mt*_ is the risk group to which an individual is assigned, and *E*_*t*_ represents the aggregate number of individuals in each infection state and includes risk group combination across the whole population (*E*_*gt*_).

The transitions between infection states are indirectly related to the decision choices *d* and non-infection state variables *k* resulting from the aforementioned decision model through assignment into risk groups *g∈ 𝒢*. Thus conditional on individual *m*’s risk group at time *t, g*_*mt*_, the model transition matrix is independent of *d*_*mt*_ and *k*_*mt*_, and only depends on aggregated risk-group level distributions of infection states. All individuals in the same risk group have the same transition matrix, so **Q**_*mt*_ = **Q**_*gt*_ for *m* ∈ *g*. Risk group assignments don’t change as a part of the infection dynamics encoded in this model, and the sum of individuals in each risk group must be constant after these transitions, until another decision occurs. The parameter vector Ψ includes all parameters for the infection model, including the way in which parameters may (or may not) be impacted by risk group membership. The infection model is a component of the overall transition matrix used in the decision model (**P**).

The epidemiological model is encoded in discrete time, with time intervals marked by the times at which we allow behavioral decisions to be updated. The time step Δ*t* can be arbitrarily small, and the rules used to generate the transition matrices **Q**_*gt*_ could themselves be a continuous model between *t* and Δ*t* (for example a system of ordinary differential equations), or a discrete model with much smaller time steps than those used in the decision model.

FIEM allows for any disease model herein the effect of decisions on disease dynamics could be translated into decision-dependent risk-group membership. In theory, the number of risk groups could be as large as the population itself, although this would dramatically increase the computational cost of the model. The rates encoded in the transmission matrix typically describe a combination of reactions that depend only on model parameters, and thus represent constant per capita rates (e.g., the rate of progressing to more advanced stages of infection typically does not depend on the prevalence of infection in the population), and and other reactions wherein the per capita transition rate depends (typically linearly) on the number of individuals in another state (e.g., in typical infection models, the rate at which individuals transition from the susceptible to infected state depends on the proportion of the population that is already infected, since they are the ones from whom transmission could occur).

#### Scenario

We designed a simple scenario based on the early part of the COVID-19 pandemic, where infection spreads in a previously unexposed population, conferring temporary immunity to reinfection, and individuals can make the decision to abstain from in-person work, reducing their contacts and risk of infection but losing income.

We track *N* individuals stratified into one of three infection states—susceptible to infection but currently uninfected (*s*), infected and infectious (capable of transmitting to others, *i*), and recovered and immune to re-infection (*r*). Thus, the set of infection states is *𝒳*= *{s, i, r}* and individual *m*’s infection state variable at a given time *t*, denoted by *x*_*mt*_, takes one of these values. Individuals who are infected sustain a utility cost *θ*_*x*_, which represents an expectation over all possible outcomes of disease (ranging from from asymptomatic infection to death). We assume that individuals have perfect information about their infection-status and that they know the aggregate distribution of infection states with a lag *l* = 7 days when making decisions for the current period.

We consider a heterogeneous population to capture and evaluate two forms of vulnerability: health and economic. Individuals are divided into two levels of socioeconomic status (SES), and two levels of vulnerability to infectious disease, both of which we assume do not change over time. No other individual-level non-infection-status state variables are tracked. Formally,

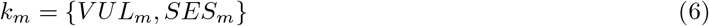

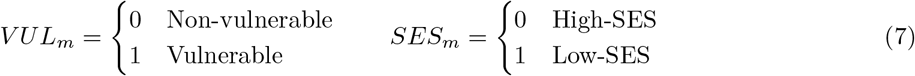

Individuals with vulnerability sustain an additional utility cost of infection, which multiples the cost by a factor *θ*_*v*_, representing their higher risk of developing more severe forms of infection.

Each period an individual makes a decision *d*_*mt*_ to work or not (we abstract from working at home):

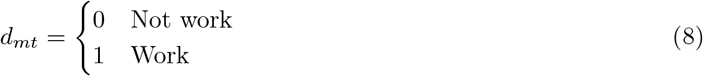

so the set of possible decisions is *𝒟*= *{*0, 1*}*.

Individuals that choose to work earn a wage (*w*_*mt*_) based on whether they are low or high SES. The amount an individual consumes, *c*_*mt*_, depends on their earnings in that period. If they work, we assume they consume their entire wages, while if they don’t work, they consume a lower “baseline” amount, which is also based on their SES status and strictly lower than their consumption if they had worked (*c*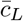 or 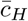 for low and high SES, respectively). Wages and consumption are formally defined as:

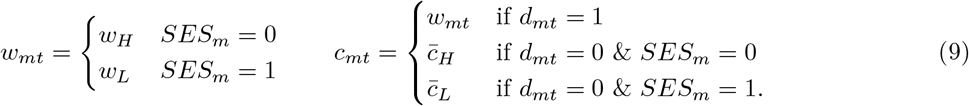

We assume there are non-monetary costs associated with working (e.g., arranging childcare, transportation, dislike of work), which we refer to as “hassle costs” and denoted by the product *θ*_*h*_*h*_*mt*_. We assume that these hassle costs follow a log normal distribution by allowing log 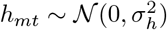. Using a distribution instead of a fixed value adds a source of inter-individual variation to the model, which while abstracting from the many sources of variation in reality, allows us to avoid all similar individuals from making the same decision under the same conditions, while still tracking a minimum number of state variables and parameters. If an individual decides to work while infected, hassle costs increase by *p*_*c*_, which could represent the discomfort of working while experiencing symptomatic disease, or stigma associated with working while visibly infectious, for example.

Under this model, an individual’s flow utility function at time *t* is specified as:

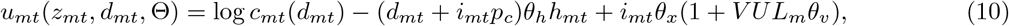

where *i*_*mt*_ is an indicator variable with value 1 if an individual is in the infected state and value 0 if they are not. We use log *c*_*mt*_(*d*_*mt*_) to consumer risk aversion to lost consumption.

Given this parameterization, the total number of risk groups is eight (vulnerability vs non-vulnerable, low- vs high-SES, and working vs not working). Since we assume that SES and vulnerability are fixed over time, the component of the state transition matrix tracking non-infection variables (**A**, is an identity matrix), and the overall transition matrix **P** is therefore defined by the infection model **Q**. We further note that, as described above, all individuals in the same risk group have the same transition matrix, thus we have **Q**_*mt*_ = **Q**_*gt*_. We describe transitions in our infection model by risk group, resulting in eight 3 *×* 3 transition matrices **P**_*gt*_.

We describe infection dynamics using a continuous-time risk-stratified SIRS (susceptible-infectious-recovered-susceptible) model. Susceptible individuals in risk group *g*_1_ can acquire infection from infected individuals in any risk group, at a rate that depends on their propensity to make contacts with individuals in corresponding risk group 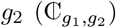 and the per-contact rate of disease transmission (*β*). Infected individuals immediately become infectious, recover from infection and lose the ability to transmit at a rate *γ*, leading to an average duration of infection of 1*/γ*. Recovered individuals are immune to re-infection, but lose immunity and become susceptible again with waning rate *α* (where 1*/α* is the average duration of protection). The model is defined by a set of possible transitions that individuals can undergo to move from one state to another, along with their corresponding rates (probabilities per time).

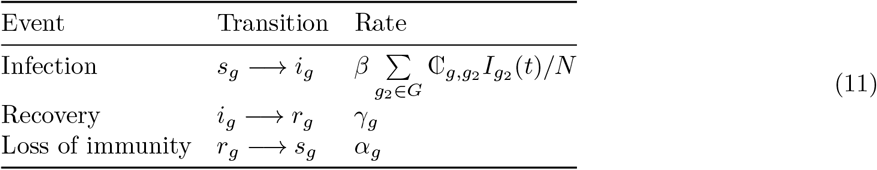

The expected value of number of individuals in each state over time under this model can be expressed as the following system of ordinary differential equations,

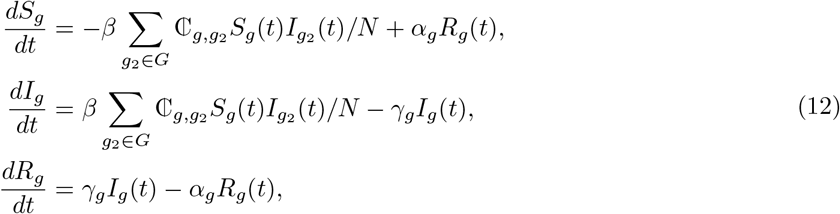

where *S*_*g*_(*t*), *I*_*g*_(*t*), and *R*_*g*_(*t*) represent the total number of susceptible, infected, and recovered individuals for each risk group at time *t*, while *N* represents the total population, computed as *N* =∑ _*g* ∈ *𝒢*_ *S*_*g*_(*t*) + *I*_*g*_(*t*) + *R*_*g*_(*t*). Risk-group membership is defined during the decision phase of the model and is fixed for each time period for which the infection model is simulated, so the total number of individuals in each risk group cannot change in the infection model.

This risk-group-level infection model is used to construct the transition matrix **P**_*gt*_ describing the probability that any particular individual in risk group *g* and infection state *x*_*t*_ at time *t* transitions to state *x*_*t*+Δ*t*_ during the current time period. The model is forward simulated between time *t* and *t* + Δ*t*, and the probability of transition is taken as the fraction of individuals who transitioned. For example,

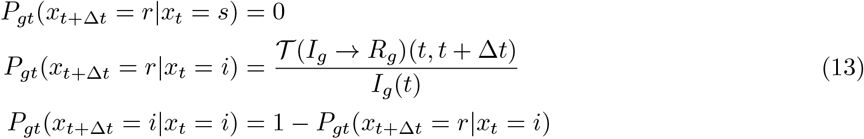

where *𝒯* (*I*_*g*_ *→ R*_*g*_)(*t, t* + Δ*t*) denotes the number of individuals transitioning from infectious to recovered in risk group *g* between time *t* and *t* + Δ*t*. Details of the numerical method used for simulating the model to future periods are provided in the “**Algorithm**” section.

Our infection model describes homogeneous mixing within each risk group and the option for heterogeneous mixing between different risk groups. Our model can incorporate heterogeneities that lead one group to have more total contacts than another group, as well as those that lead to preferential (i.e., assortative) mixing between certain groups. The way in which the decision to work—via risk group membership—influences disease risk is through the form of the contact matrix 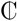. We assume there is a baseline number of contacts each individuals in the population would have if the entire population was high-SES and non-working,*C*_0_, which would result in a baseline probability of contacting any other individual in the population of *C*_0_*/N*. For heterogeneous populations, we allow contact propensities to be group-dependent. For individuals who work, we let the propensity to contact other working individuals be increased by a factor *σ*_work_. Low-SES individuals have a *σ*_SES_ increase in the probability of contact with others with low SES. We assume there is some degree of preferential mixing by vulnerability and risk group—regardless of the decision to work—so that the propensity for individuals to contact risk groups with the same vulnerability and SES is increased by *σ*_1_. Mathematically, the contact propensities are,

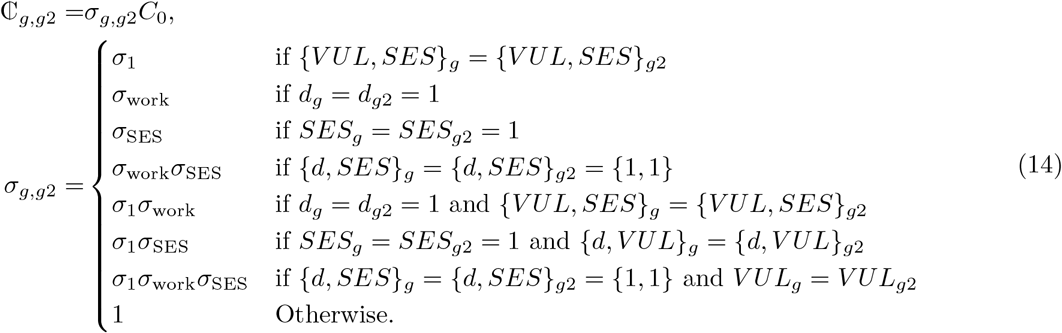

Or, written as a matrix,

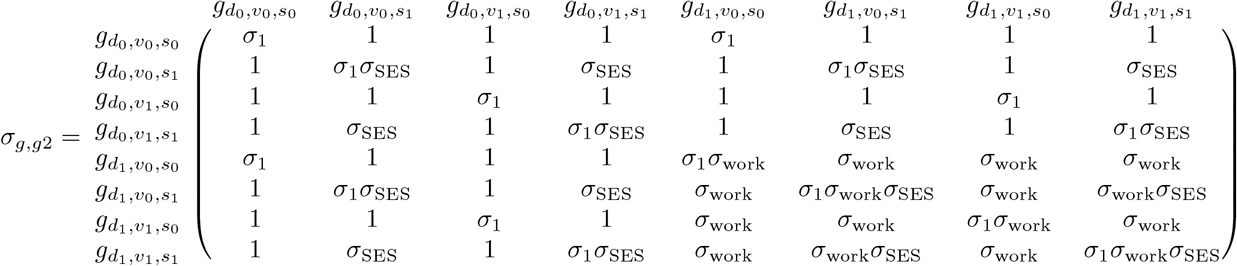

where the subscripts *d*_0_ and *d*_1_ are used to denote risk group members do not and respectively do work, *v*_0_ and *v*_1_ are used to denote risk group members are not or are vulnerable, and *s*_0_ and *s*_1_ are used to denote members are of high or low SES.

Note that we our formulation of the force of infection term in Eqs (11) and (12) is intentionally somewhere between the traditional density-dependent and frequency-dependent assumptions (Begon et al., 2002). We wanted transmission to be invariant to the total population size *N*, but to react to the distribution of individuals across risk groups (*N*_*g*_), since risk group size changes dynamically in our model, unlike commonly used age- and spatial structures in infectious disease models which are more rigid. For example, when fewer individuals choose to go to work, the remaining working individuals contact fewer other individuals in their workplace. Hence, our formulation retains the relative probability of contacts between risk groups, but not the absolute number of contacts. The probability an individual in group *g* contacts someone else in group 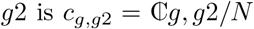 and the *number* of contacts an individual in group *g* has with someone in group *g*2 is 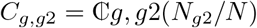.

#### Parameters

Our model includes parameters that characterize the decision model (Θ) and infection model (Ψ). To showcase FIEM, we chose parameter values for the model to reasonably capture the trade-off between health and wealth that individuals faced during the early stages of the COVID-19 pandemic. Parameter values were either fixed based on values estimated in the literature, or chosen to produce realistic model output. In future work, parameter values could be more formally inferred using a combination of datasets describing infection dynamics, preventative behaviors, labor participation, etc.

The parameters of the decision model describe how an individual’s utility depends on their state variables and the decision they make (Equation 10). These parameters are listed, along with descriptions, interpretations, assumed values, and data sources, in Table 2. A key way to validate our choice of parameter values is to show they are broadly consistent with estimated cost of COVID-19 infection in dollar terms. We obtain estimates for the monetary value per statistical case (VSC) of COVID-19 from the United States Department of Health and Human Services (Kearsley, 2024). They report VSCs of $13 million per fatality, $1 million for severe to critical cases, and $5,000 for mild cases. Given an estimated COVID-19 infection fatality risk of 0.068% (Levin et al., 2020) and an infection hospitalization risk of 2.6% (Griffin et al., 2024) for individuals 35–44 years old, the implied average VSC is $41,387. With an average duration of seven days, this value translates to an estimated economic burden of $5,912 per day of infection. By calculating the average compensating variation in our simulated population between being in the infected state vs. not being in the infected state (i.e., the amount of consumption that would leave an individual indifferent in utility terms between the two options), we extracted the daily cost of infection implied by our model parameterization to be $5,970.

To ease the computational burden associated with having to uniquely evaluate value functions for each individual, we discretize distribution of hassle costs *h* into three categories:

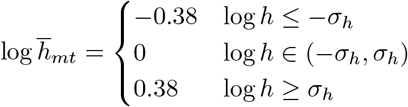

where the value for each category is approximately the conditional mean of log *h* within each interval for a distribution standard deviation *σ*_*h*_ = 0.25. Together with the utility parameter *θ*_*h*_, we set the baseline hassle cost for the average individual at $116 per day. A one standard deviation increase results in a hassle cost of $193 per day, while a one standard deviation decrease brings it down to $73 per day. This range captures the variation in hassle costs across individuals, reflecting different levels of inconvenience or burden.

The parameters of the infection model are summarized in Table 3. We make the simplifying assumption that risk group membership affects *only* contact probabilities, and does not confer any differential susceptibility, duration of infectiousness, or duration of immunity (i.e., *γ*_*g*_ = *γ* and *α*_*g*_ = *α*). However, this assumption can easily be relaxed to allow for additional complexity or to model different decisions in future work.

We assume an average duration of infectiousness of 7 days, based on studies estimating generation intervals, duration of viral shedding, and symptom duration, which corresponds to an average rate of recovery from infectiousness of *γ* = 1/7 = 0.14 days (Byrne et al., 2020; Ali et al., 2020; Hakki et al., 2022; Lavezzo et al., 2020). After recovering from infection, we assume immunity that confers perfect but temporary protection against re-infection, with the average duration of protection of 6 months (rate of waning *ω* = 0.004/day). This value was taken from two meta-analyses that measured the efficacy of protection against the Omicron variant as a function of time since prior infection with pre-Omicron variants. The rate of waning of protection against pre-Omicron variants was estimated to be significantly slower, but is less relevant due to the emergence of the Omicron variant in November 2020. Since the decay of protection is assumed to be constant in our simplified model but observed to be variable over time in these studies, we chose this waning rate to be consistent with the residual protection remaining at 7.5 months (the time between peak pre-Omicron infections in spring 2020 and peak Omicron infection in late 2020/early 2021 in many regions of the world) (Stein et al., 2023; Bobrovitz et al., 2023).

We assume a baseline rate number of contacts at any given time *C*_0_ = 4, which is informed by the average effective number of contacts occurring at home relevant to respiratory disease transmission(Mistry et al., 2021; Mossong et al., 2008; Prem et al., 2017). To model variation in the propensity of contact between risk groups, we assume individuals who work have 4 times the probability of contact with other individuals who work (*σ*_work_ = 4), which is roughly based on observed numbers of average work contacts and our assumption that 75% of the population is working at baseline. We additionally assume individuals who are of low socioeconomic status have 1.5 times the probability of contact with other low socioeconomic status individuals (*σ*_SES_ = 1.5), based on numerous lines of evidence suggesting low SES status is associated with more household crowding as well as jobs that require more high-risk in-person interactions (Papageorge et al., 2021; Dingel and Neiman, 2020; Nande et al., 2021; Jay et al., 2020; Kamis et al., 2021; Emeruwa et al., 2020). Finally, to add an additional source of assortitivity (“homophily”) to capture the many ways in which human interactions tend to include preferential mixing across many sociodemographic and behavioral characteristics (McPherson et al., 2001; Newman, 2002; Smith et al., 2014; Hilman et al., 2022), we assume individuals have 1.5 times greater probability of contact with individuals belonging to the same vulnerability and SES groups (*σ*_1_ = 1.5). Note that because our model is based on proportional mixing and is does not include a formal network model of discrete contacts, only the product of the transmission rate *β* (which is calibrated to give the desired epidemic growth) and the contact matrix 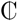 influences results, not their separate values nor the exact number of contacts they imply.

#### Algorithm

In this section we describe how to recover the solution of the combined behavior-disease model each period. To summarize, at the beginning of a period, each individual is associated with state variables *z*_*mt*_ = *{k*_*mt*_, *x*_*mt*_, *E*_*t*_*}*. An iterative numerical procedure (Algorithm 1), described in detail below, is used to numerically recover the net-of-errors value functions (Equation 4) for each possible combination of state variables and decisions. These are then used to calculate the conditional probabilities of making different decisions for each individual (Equation 5), which in turn allow us to construct the model’s risk groups. In the intervening time period Δ*t* until the next decision is made, infection levels evolve in a risk-group specific manner according to the infection dynamics model. The cumulative transitions in infection status during the proceeding period are used to construct the transition matrix describing expectations about changes in infection states in the future, that then feeds into the estimated value functions—and hence decisions—at the next decision update. Continually repeating this procedure of successively approximating the value functions, calculating decision probabilities, and executing changes in decision status and infection levels for the next time step allows us to simulate the paths of choices individuals make over time as their state variables evolve.

We use the nested fixed point method, a type of successive approximation introduced by Rust (1987), to recover the full solution to an individual’s dynamic discrete choice problem at each time step. To understand how this method works, consider the following simple example. Suppose the decisions *d* and states *z* are binary so there are four combinations. We use *a* and *b* to index the states *z*, and 0 and 1 to index the decisions *d*. Without loss of generality, transitions between *a* and *b* are captured by the transition matrix **P**_*mt*_(*d, z*; Ψ) (note since we are here describing decision making at the individual level, we index by *m*). This process produces a system of equations based on the net-of-error value functions. To slightly abuse notation, let 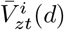 denote iteration *i* of the net-of-error value function for an individual with state variables *z* making decision *d* in period *t*:

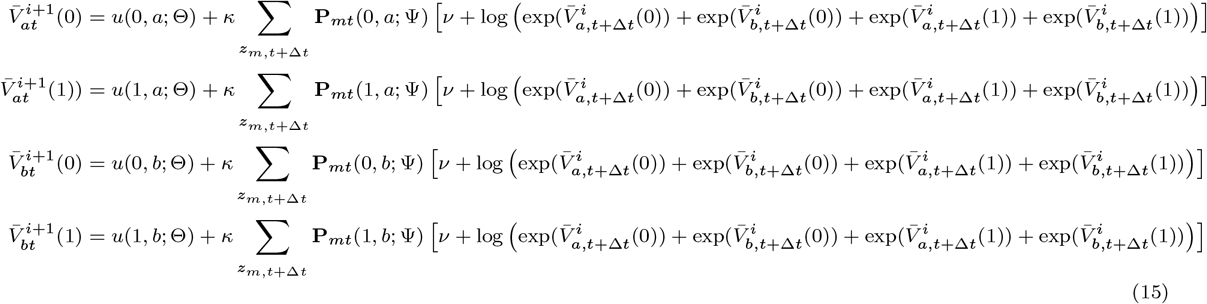

Rust (1987) proved that under certain assumptions (i.e., *κ <* 1 and bounded flow utility), the true value function is a fixed point of this system of equations. Thus, given an initial guess for the values of 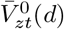, we can iterate on this system to recover the numerical values for the value functions within a pre-specified tolerance for a given utility parameter vector Θ.

The infectious disease dynamics are described by continuous rates (Equation 11) at which individuals in a given risk group transition between susceptible (*S*(*t*)), infected (*I*(*t*)), and recovered (*R*(*t*)) states. Although we model disease dynamics on a continuous time scale, the decision model requires a transition matrix describing infection dynamics over a discrete time period. To efficiently make this bridge in timescales while avoiding numerical errors with discretizing continuous models, we calculate updates of the infection model in smaller time steps, *δt*, using a simple discrete time Euler update. For the SIRS model described in Equation (12), the transition probabilities are

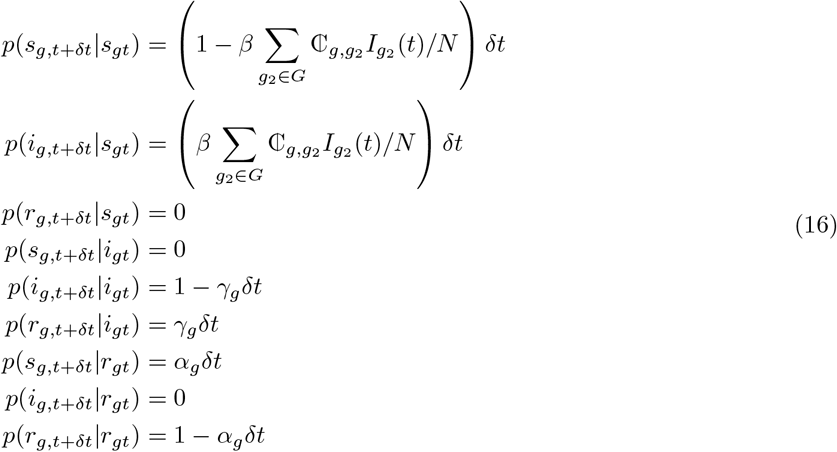

which are the elements of the transition matrix **P**_*gt*_. To update the state of each individual, we randomly choose a transition to occur using a multinomial distribution with parameters given by the transition probabilities.

We conclude with brief comments on computation and tractability. As the model includes more state variables or decisions, the number of components within equation (15) increases, leading to higher computational burden. Thus it is possible to specify a model that is so rich that solving for the fixed point of equation (15) becomes intractable. However, we do not see this as a major concern to the FIEM. The computational burden can be alleviated by using parallel computing (i.e., if your parameterization allows you to break up the net-of-error value functions into groups) or through the use of fix point acceleration algorithms. Additionally, computational resources are still continuously improving. Finally, an effective model should be parsimonious enough to answer the question at hand without introducing unnecessary features that can exacerbate any tractability issues.

##### Algorithm 1

FIEM Solution

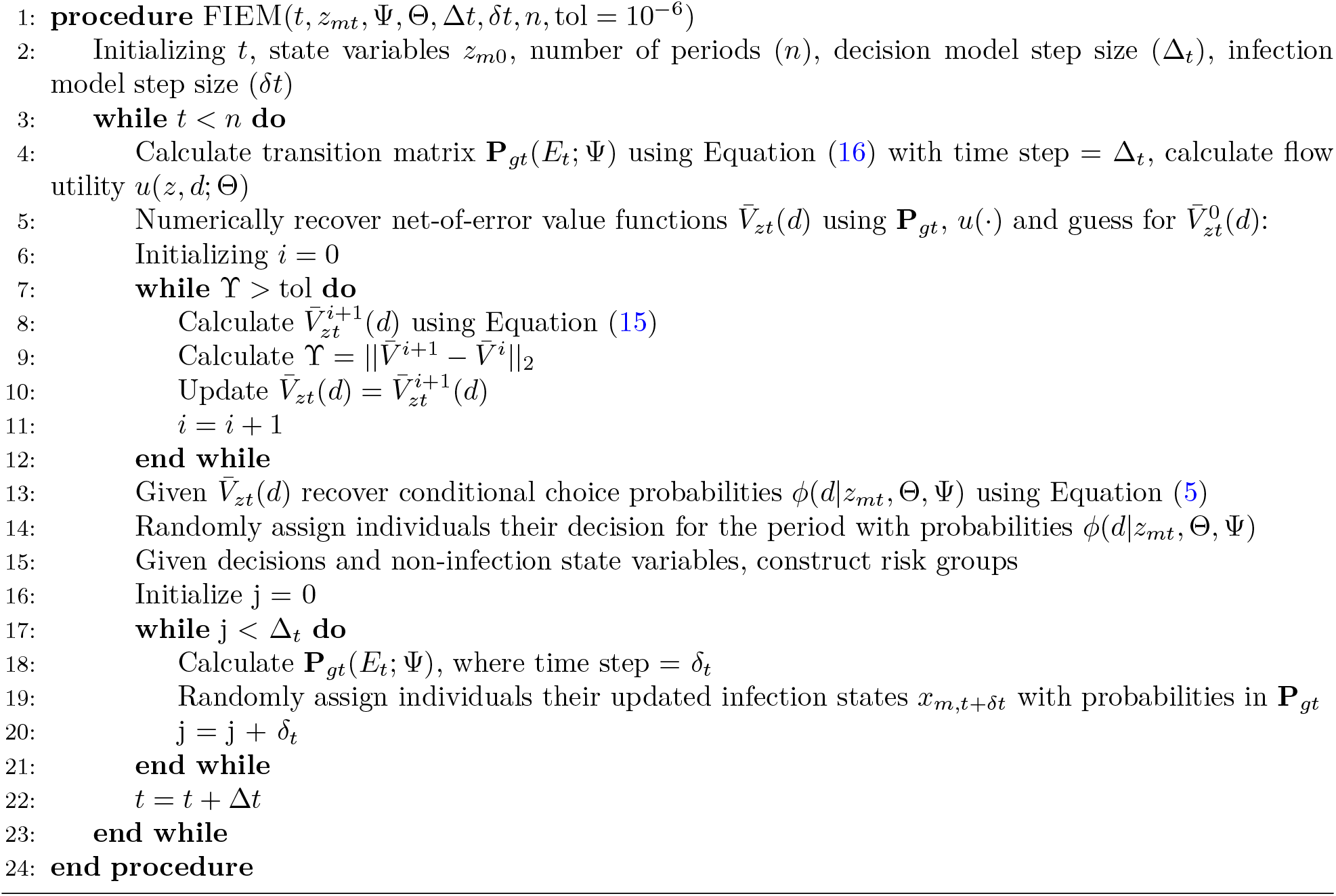

#### Policy Scenarios

In the main text we use FIEM to simulate the effects of four alternative policies on disease spread, labor supply, and other individual outcomes. The motivation behind each policy is to reduce the number of contacts individuals have within a period. If individuals have fewer contacts then the infectious disease will not spread as much through the population absent the intervention. The policies differ in terms of the mechanism that is used to reduce contacts, which groups are targeted, and their fiscal costs to the government. Here we detail how these policies are implemented as part of the FIEM framework. Individuals cannot anticipate any of these policies and do not modify their behavior in the run up to their implementation.

The first policy is the labor restriction. This policy loosely resembles some of the so-called “lockdown” measures used during the COVID-19 pandemic that resulted in some businesses or agencies not being open. Under this policy, a randomly chosen share of the population is forced to remain at home, while the other portion of the population can choose to work or not. In practice this means that the portion of the population that is under the restriction does not solve the dynamic programming problem because they are only able to take a single action each period. In terms of the notion introduced previously, this policy reduces the set *𝒟* for the randomly chosen portion of the population to a singleton (i.e., not working for the period).

The second policy is an unconditional cash transfer. This policy also resembles actions taken during the recent COVID-19 pandemic when governments provided direct cash payments to individuals. Each period, individuals receive a cash transfer from the government. This transfer is added to the income the individuals receives that period based whether they work or not (i.e.,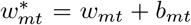, where *b*_*mt*_ is the value of the transfer provided). Since we rule out savings, these cash transfers are consumed by individuals that period. The third policy is a conditional cash transfer. It is the same as the previous policy but only individuals that choose not to work will receive the payment from the government (i.e., individual *m* will only receive the payment *b*_*mt*_ if 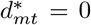). These policies were used later on in the COVID-19 pandemic. Individuals know they will receive the government payment if they choose not to work that period.

The final policy simulates the effects of paid sick leave. While this policy was not part of the response to the COVID-19 pandemic used by most governments, it is designed based the same core motivations. It seeks to directly target the populations that present the greatest risk of spreading the disease through the population by giving them a direct incentive to reduce contacts. This incentive arises through the same channel as the cash transfer measures by increasing the income and consumption of targeted individuals. The transfer is only available to individuals that are infected that choose not to work. Once the policy is activated, individuals are aware that they can receive this payment if they become infected and choose not to work. We do not assume there are any barriers to taking up the benefit.

#### Model Validation

In this section we present two sets of validation exercises. The first type of validation experiments focuses on ensuring the model of individual behavior responds in predictable ways, while the second type performs a similar assessment of the epidemiological model. The value of these exercises is that they confirm the models of individual behavior and epidemiological disease spread are designed and implemented correctly. They also develop intuition for what factors drive behavior and disease spread within each component of our framework in isolation. Establishing this baseline is important for evaluating the predictions and performance of the integrated framework where both models feedback to each other.

#### Infection model validation

In this section we perform an additional set of validation exercises for the risk-stratified disease spread model. We explore the behavior of the disease dynamics when individuals’ decisions do not change in response to the overall dynamics. We consider the disease dynamics when risk group membership stays constant (Figure S1). This is equivalent to a standard compartmental disease model with some introduced level of heterogeneity in contact patterns.

First, we note that when no individuals work, the disease is eliminated, as there is insufficient (or no) contact required to sustain an epidemic. Conversely, when all of the population work, the epidemic is quick and sharp. When an intermediate proportion of individuals work, e.g. half the population goes to work, the peak of the epidemic is significantly reduced. The decrease in the population working corresponds to a decrease in contact propensity, thus lowering the basic reproductive number.

We also consider the effect of different proportions of other static state variables (vulnerability and socioeconomic status). All other states constant (socioeconomic status and working state), vulnerability does not affect disease dynamics when there is no dynamic decision making; this is an expected result when vulnerability only feeds back to decision making, and it is the decision to work that inevitably drives disease dynamics.

Similarly, when the decision to work is constant, higher ratios of low- to high-socioeconomic status also increases the peak size and initial exponential growth rate of the epidemic. We note an important behavior, in that these dynamics hold true as a result of our parametrization of our contact matrix, where individuals have a higher probability of contact with those in their own risk group, workers interact with other workers more often, and low socioeconomic status individuals also interact at a higher propensity to contact other low-socioeconomic status individuals.

We also note that due to the choice of our SIRS compartmental structure, disease equilibria can result in oscillations, as the population cycles between susceptibility and recovery.

#### Behavioral model validation

To test the calibration and specification of our model of individual behavior we perform four sensitivity analyses. Each of these exercises should produce a predictable response from the behavior model. Should our model replicate these expected responses, then we can be confident it is specified and implemented correctly.

First, we shutdown the disease component of the model and assess how the decision to work responds to the returns of working. Under this setup, the returns to working are captured by the relative difference in the amount an individual consumes (captured by the utility function) if they work vs not work. Since this experiment shuts down other factors that determine utility, we should see the probability an individual decides to work increases as the relative difference in the utility from working increases. This pattern is exactly what we see in Figure S2 panel (a), which plots the probability an individual works as a function of the difference in utility between working and not working.

Next we introduce the infectious disease dynamics into the model. For this specification the probability of getting infected is independent of the decision to work or not. Individuals that do get sick pay a utility cost while they are infected. Individuals still earn utility from consumption, which depends on their decision to work. Given this set up we should expect two patterns to emerge. First, conditional on an individual’s health state (i.e., susceptible or infected) the probability of working or not should depend on the relative gap in the payoffs between both actions. This feature is driven by the independence of the infection probability and the labor supply decision. Second, being infected should act like a “fixed cost” in utility terms.

To assess whether the model captures these features in Figure S2 panel (b) we plot the value functions by infection state and labor supply decision as a function of the utility cost of infection. There are two takeaways. First, as the utility cost of infection increases the gap between the value functions for susceptible and infected individuals, consistent with the fixed cost of infection we expected. Second, conditional on a utility cost of infection and a health state, the gap between the value functions for working and not working is constant, which is consistent with probability of infection being the same for both individual behaviors.

The next experiment takes the prior one and allows the disease to impact the utility from working. Specifically, in addition to receiving utility from consumption, individuals that choose to work while infected pay an additional utility cost. Given this additional feature we should expect to see a response in the utility and corresponding behavior of infected individuals as the additional cost of working while infected increases. Panel (c) of Figure S2 reports the value functions for susceptible and infected individuals if they choose to work or not work as a function of the additional cost of working if infected. The value functions for susceptible and infected individuals that do not work are parallel, which reflects the gains from working and the penalty associated with being infected. The value function for infected individuals that choose to work declines as their penalty of working increases. As shown in Panel (d) of Figure S2, these declines in the value function of working while infected translates into sharp decreases in the probability that an infected individual chooses to work. Moreover this change does not alter the probability that susceptible individuals choose to work within the model.

From here we investigate how the model reacts when allowing the probability of getting infected depends on the decision to work or not. To isolate the effect of this feature we shutdown the additional cost of working for infected individuals that we introduced in the previous experiment. We should expect to see shifts in behaviors as the probability of infection conditional on work decision gets larger, as this will reflect individuals are rationally responding to engage in the behaviors that are less likely to get them infected and achieve greater utility. Figure S3 plots a heatmap of the probability a susceptible individual chooses to work as a function of the probability of getting infected if they work and not work. We note two takeaways. First, when facing the same probability of getting infected, the probability of working or not is equal. Second, as the probability of getting infected if an individual chooses to work gets larger, the probability of choosing to work falls. Related, as the probability of getting infected while not working increases, the probability an individual chooses to work rises.

Each of these exercises produced the anticipated effects. In particular, they demonstrate how individual behavior responds to its immediate payoffs as well as payoffs that materialize in later periods. These components are important for our integrated framework where individual decisions today impact aggregate disease dynamics for the next period, which in turn influence behavior.

## Data Availability

No data was produced in the present study. Code used to simulate the model is publicly available in on Github.

https://github.com/HopkinsIDD/epi-econ

## Supplementary Information

### Supplementary Tables

**Table S1:**
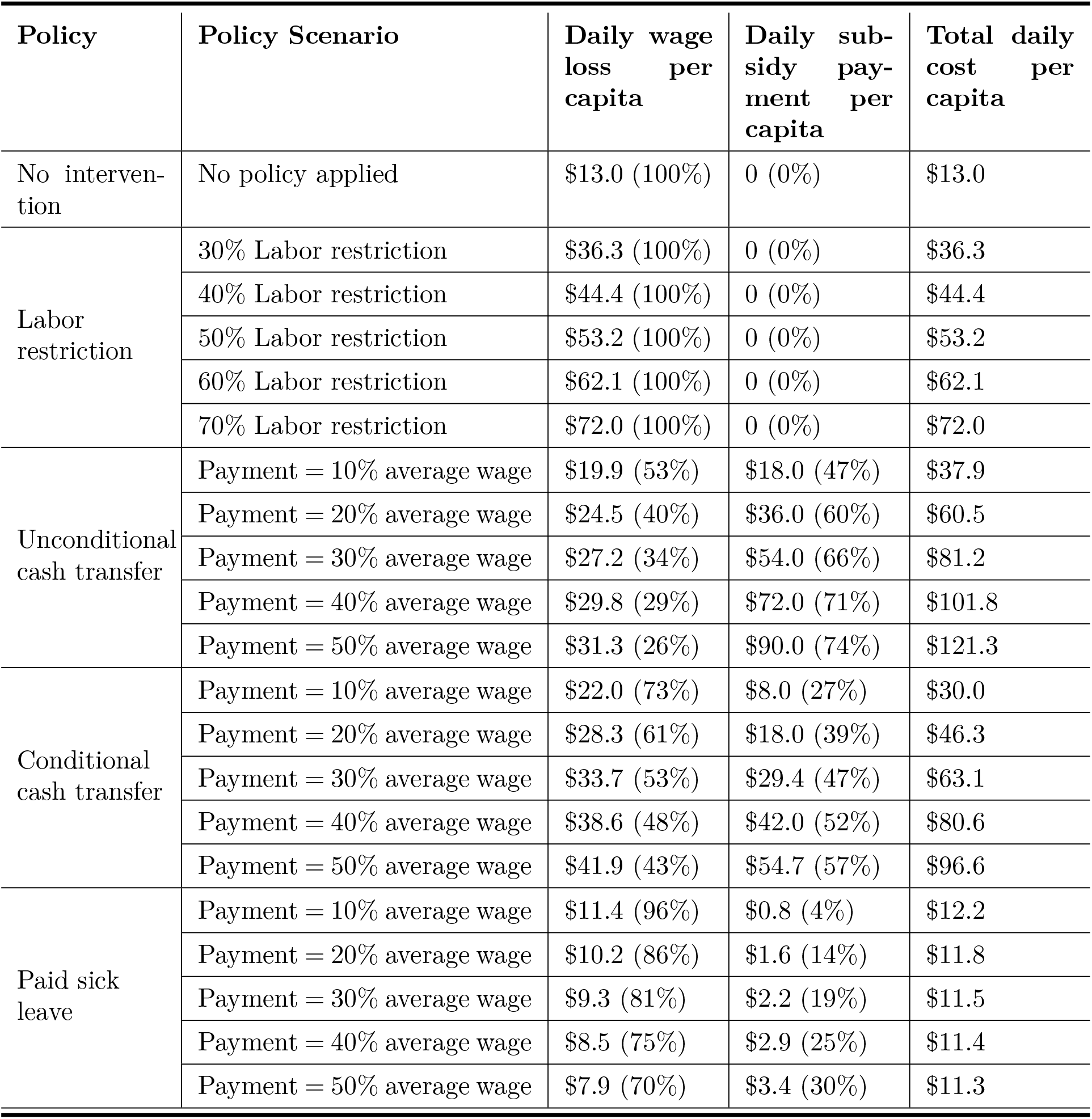
A summary of policy scenarios and resulting cost breakdown by income loss and subsidy payment. Values in parentheses indicate the proportion of each cost component to the total cost. Total daily cost per capita includes both lost wages due to the disease (compared to a disease-free scenario with 75% labor participation) and the cost of any subsidy payments provided.

| Policy | Policy Scenario | Daily wage loss per capita | Daily subsidy payment per capita | Total daily cost per capita |
| --- | --- | --- | --- | --- |
| No intervention | No policy applied | \$13.0 (100%) | 0 (0%) | \$13.0 |
| Labor restriction | 30% Labor restriction | \$36.3 (100%) | 0 (0%) | \$36.3 |
| | 40% Labor restriction | \$44.4 (100%) | 0 (0%) | \$44.4 |
| | 50% Labor restriction | \$53.2 (100%) | 0 (0%) | \$53.2 |
| | 60% Labor restriction | \$62.1 (100%) | 0 (0%) | \$62.1 |
| | 70% Labor restriction | \$72.0 (100%) | 0 (0%) | \$72.0 |
| Unconditional cash transfer | Payment = 10% average wage | \$19.9 (53%) | \$18.0 (47%) | \$37.9 |
| | Payment = 20% average wage | \$24.5 (40%) | \$36.0 (60%) | \$60.5 |
| | Payment = 30% average wage | \$27.2 (34%) | \$54.0 (66%) | \$81.2 |
| | Payment = 40% average wage | \$29.8 (29%) | \$72.0 (71%) | \$101.8 |
| | Payment = 50% average wage | \$31.3 (26%) | \$90.0 (74%) | \$121.3 |
| Conditional cash transfer | Payment = 10% average wage | \$22.0 (73%) | \$8.0 (27%) | \$30.0 |
| | Payment = 20% average wage | \$28.3 (61%) | \$18.0 (39%) | \$46.3 |
| | Payment = 30% average wage | \$33.7 (53%) | \$29.4 (47%) | \$63.1 |
| | Payment = 40% average wage | \$38.6 (48%) | \$42.0 (52%) | \$80.6 |
| | Payment = 50% average wage | \$41.9 (43%) | \$54.7 (57%) | \$96.6 |
| Paid sick leave | Payment = 10% average wage | \$11.4 (96%) | \$0.8 (4%) | \$12.2 |
| | Payment = 20% average wage | \$10.2 (86%) | \$1.6 (14%) | \$11.8 |
| | Payment = 30% average wage | \$9.3 (81%) | \$2.2 (19%) | \$11.5 |
| | Payment = 40% average wage | \$8.5 (75%) | \$2.9 (25%) | \$11.4 |
| | Payment = 50% average wage | \$7.9 (70%) | \$3.4 (30%) | \$11.3 |

### Supplemental Figures

**Figure S1:**
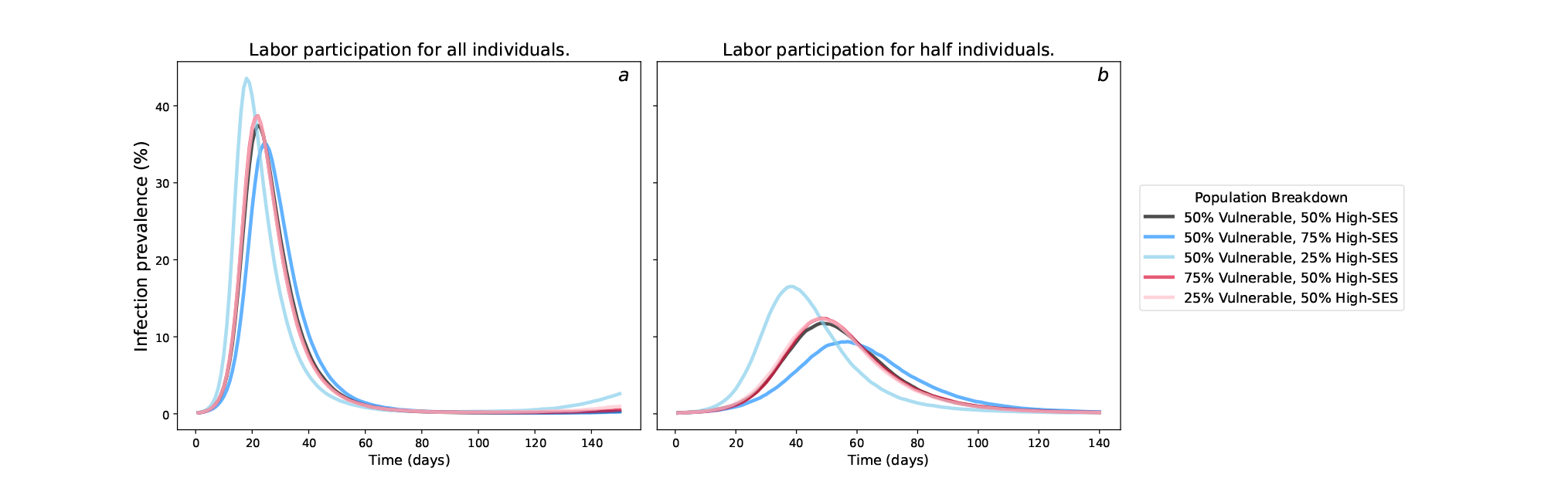
Model testing with fixed labor participation. This figure shows the infection prevalence (%) under two fixed labor participation scenarios: (a) infection prevalence when all individuals are participating in the workforce, and (b) infection prevalence when only half of the individuals are participating. The color scale represents different initial population distributions with varying heterogeneous group sizes.

**Figure S2:**
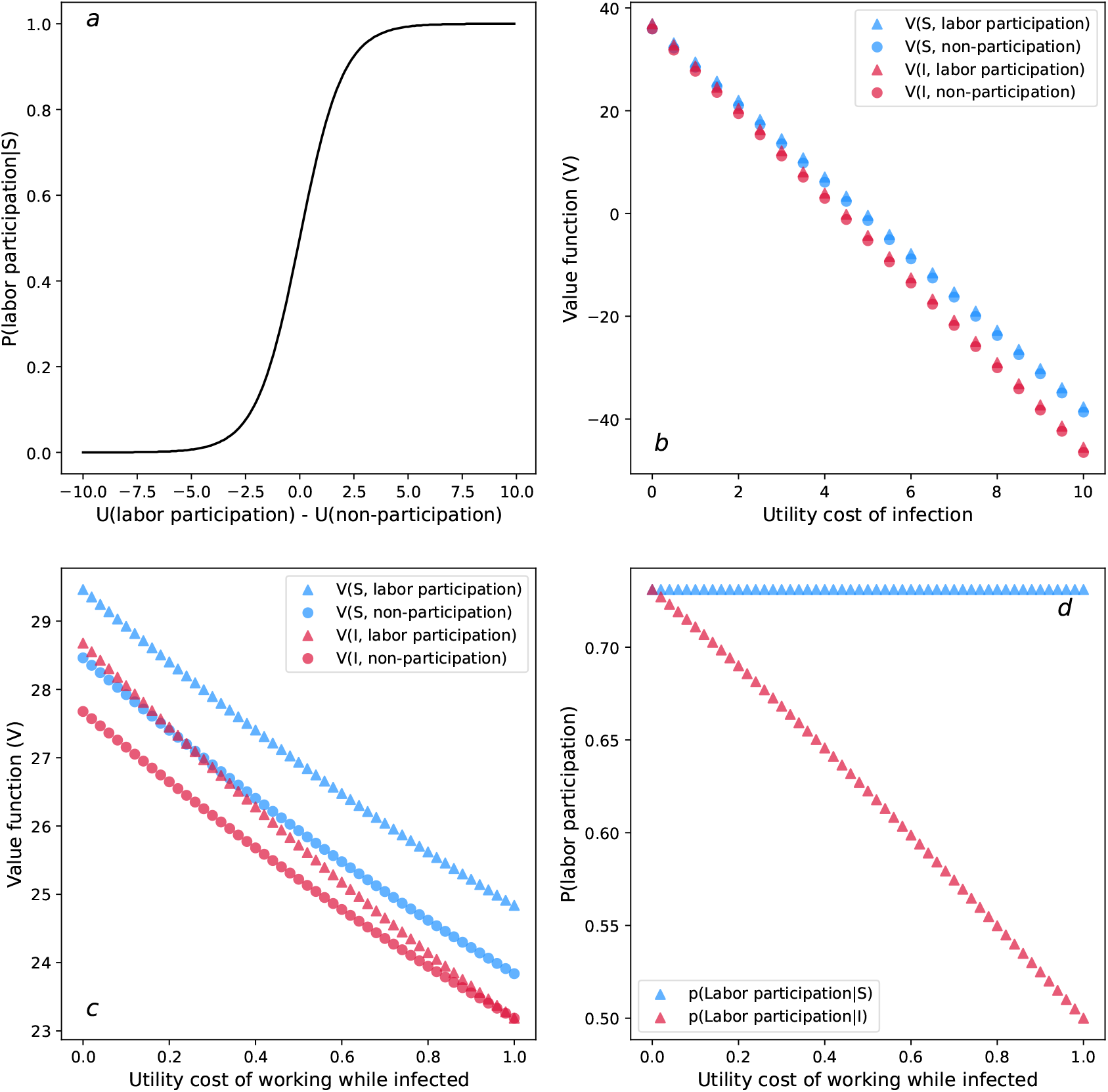
Model calibration for individual labor participation behaviors. This figure analyzes labor participation under four distinct scenarios: (a) Baseline scenario without disease dynamics: Assess how labor participation decisions respond to the utility difference between labor participation and non-participation without considering the impact of disease. (b) Disease with fixed infection probability: Assume a constant probability of infection, independent of individual behavior. The plot illustrates the value functions of infection state and labor participation decision as a function of the utility cost of infection. (c) Infection impacts utility from working: Individuals working while infected face an additional utility cost beyond the utility from consumption. The plot shows value functions by infection state and labor participation decision as a function of the utility cost of infection. (d) Labor participation vs. utility cost of working while infected: Under the same conditions as in (c), this plot depicts labor participation rates as a function of the utility cost of working while infected.

**Figure S3:**
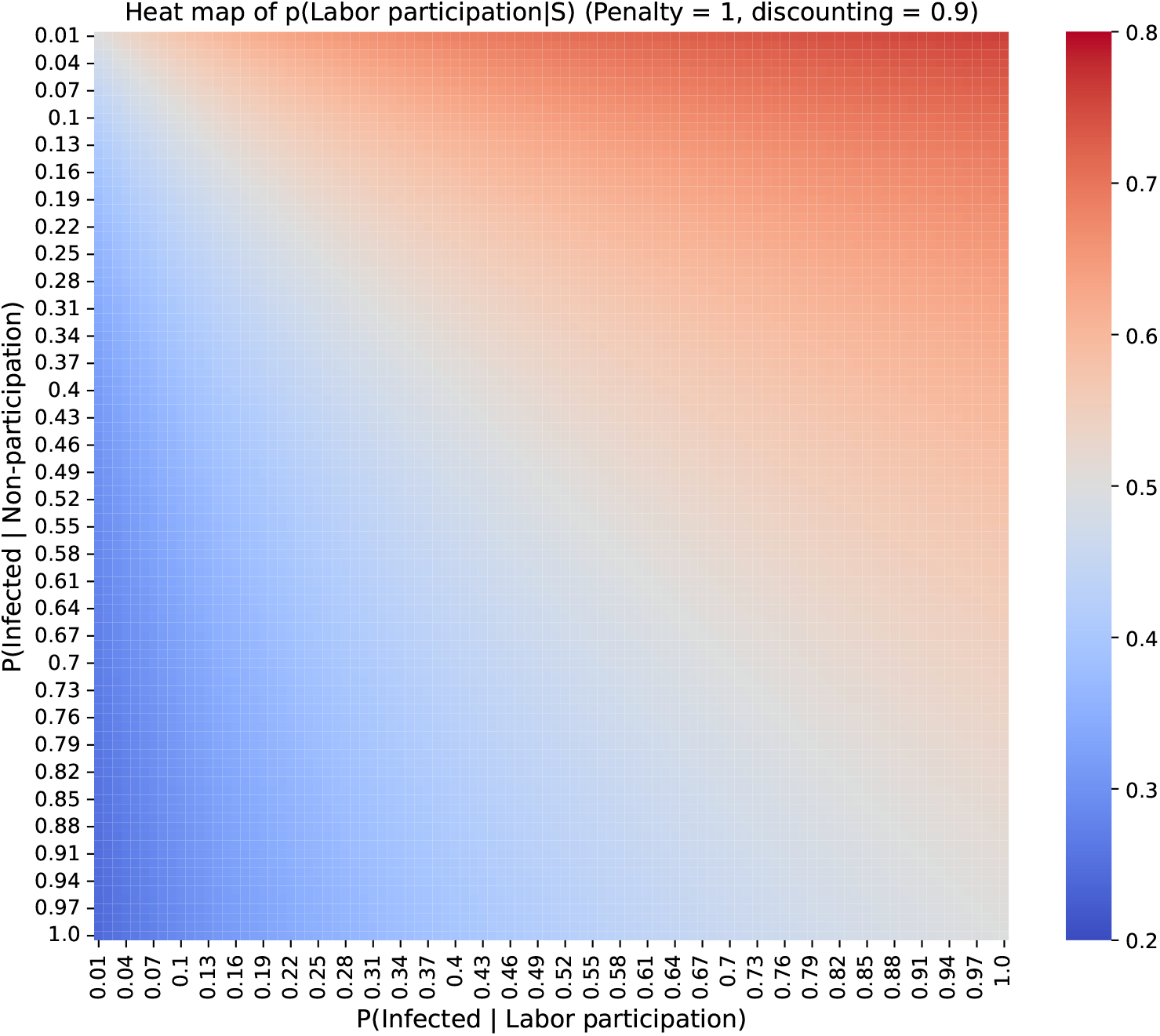
Heatmap of labor participation decisions based on infection probability. The x-axis represents the probability of getting infected while participating in the workforce, and the y-axis represents the probability of getting infected while not participating. The color scale indicates the probability of choosing labor participation, with redder colors denoting a higher likelihood of participation.

**Figure S4:**
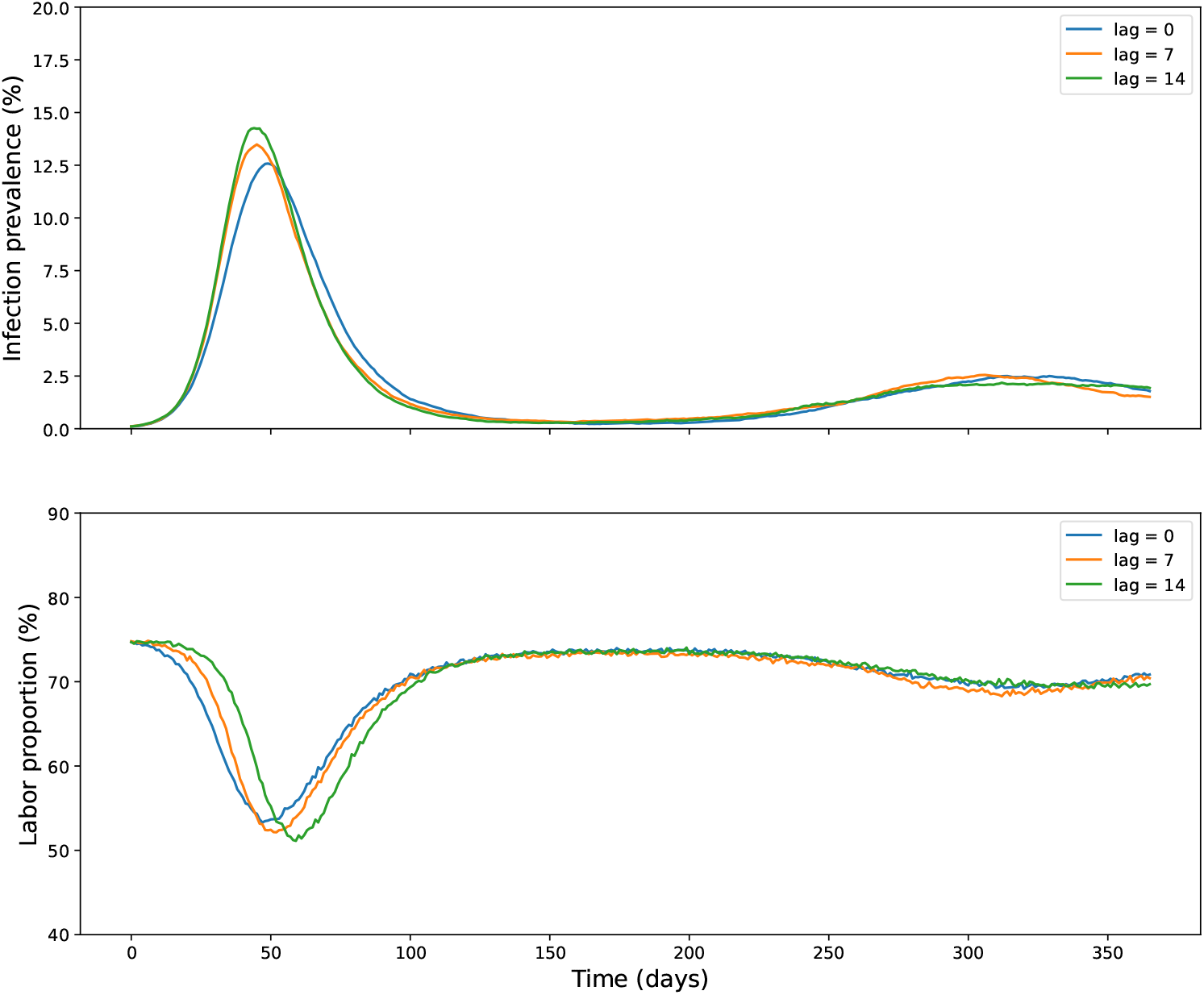
Sensitivity analysis to the amount of information individuals have about the aggregate infection rates when making decisions. A lag of 0 means individuals know the true aggregate infection rate in real time, a lag of 7 means individuals know the aggregate infection rate from 7 periods prior to the current one, and a lag of 14 means individuals know the aggregate infection rate 14 periods prior to the current one. The top panel reports the share of the population that is infected and the bottom panel reports the share of the population that works under these three information structures.

**Figure S5:**
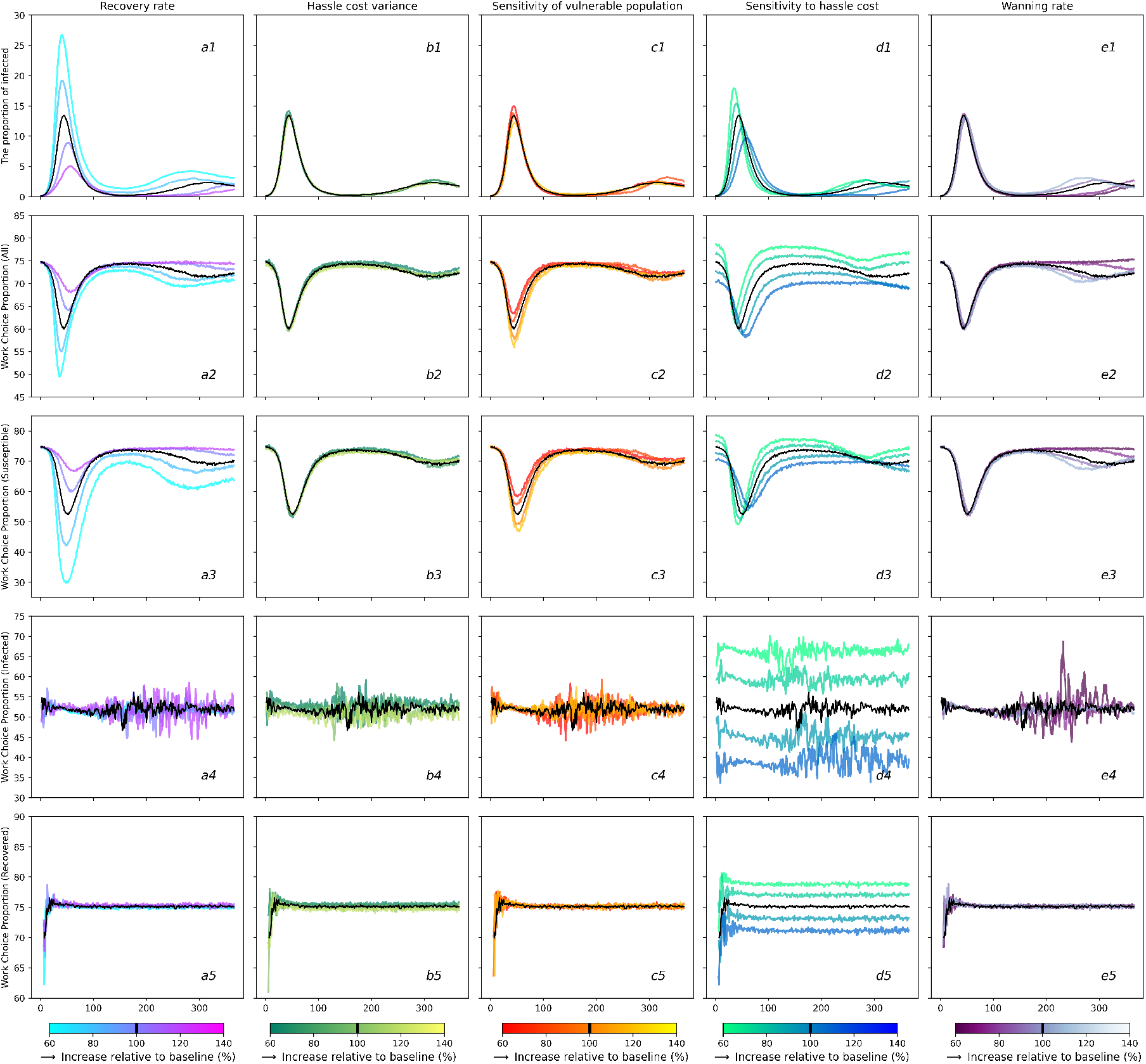
Sensitivity analysis of selected model parameters. The black curves in each panel show our baseline parameter values as given in Tables 2, 3, and **??**. Row (1) reports the population infection rate, Row (2) reports the share of the population that choose to work, and rows (3–5) report the work decisions by susceptible, infected and recovered groups. Column (a) varies the recovery rate, (b) alters the variance of hassle cost, (c) varies the sensitivity of vulnerable populations to the infection risk, (d) alters the sensitivity of hassle cost, and (e) varies the immunity winning rate

**Figure S6:**
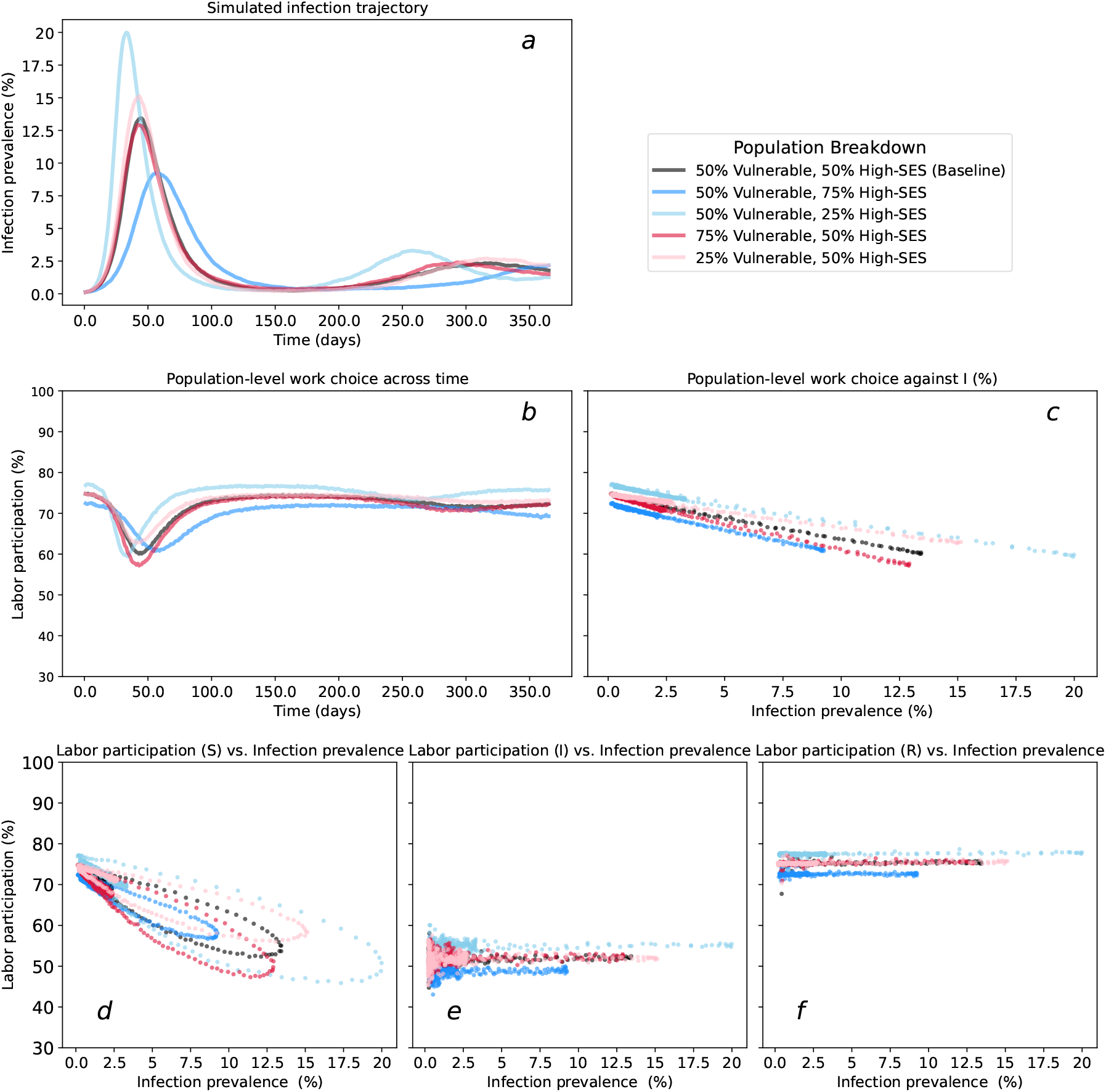
Illustration of decision dynamics with heterogeneous populations. Panel (a) captures the share of the population that is infected; (b) includes the share of the population that choose to work; (c) shows the population infection rate against the share of the population that choose to work; (d–f) show the work choice probabilities for susceptible, infected, and recovered health state groups respectively. Five populations are simulated and differ based on the distribution of non-infection characteristics (i.e., vulnerability to the disease and SES). The black curve represents the baseline parameter values.

**Figure S7:**
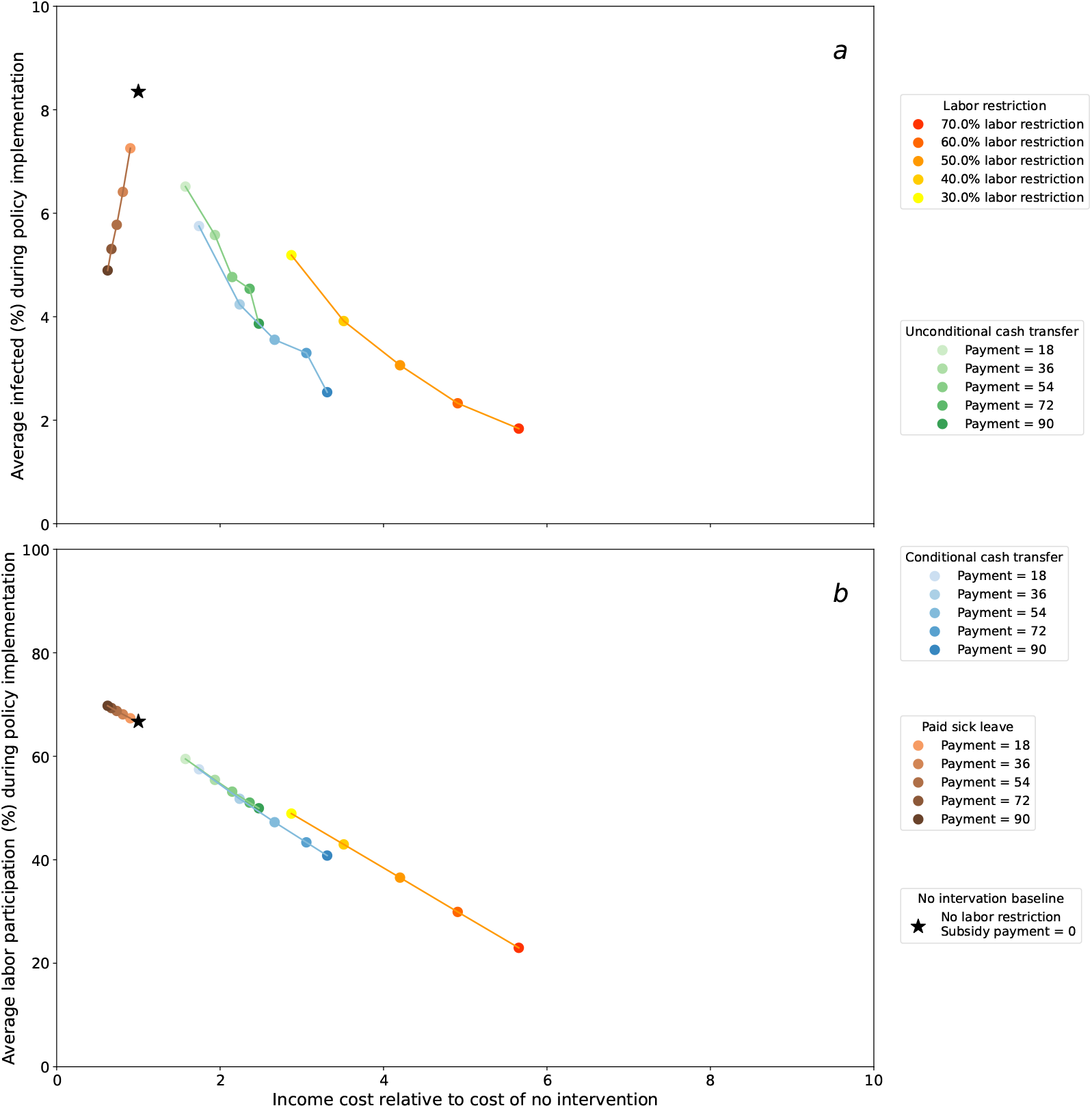
Average infection and labor supply by different policy interventions. This figure plots the average share of the population that is infected and working under different policy interventions as a function of their income loss. Panel a plots the average infection rates over the time period when the policies are in effect, while panel b plots the average share of the population that chose to work during this time period. The cost captures lost wages an individual experiences relative to their baseline wage earnings predicted by the model when there is no intervention. The simulated policies include: a labor restriction policy that limits how much of the population is able to choose to work; an unconditional cash transfer that is delivered to all individuals each period; a conditional cash transfer that is delivered to individuals that choose to not work each period; and a paid sick leave policy, which allows infected individuals to earn their full wage if they choose to not work while infected.

**Figure S8:**
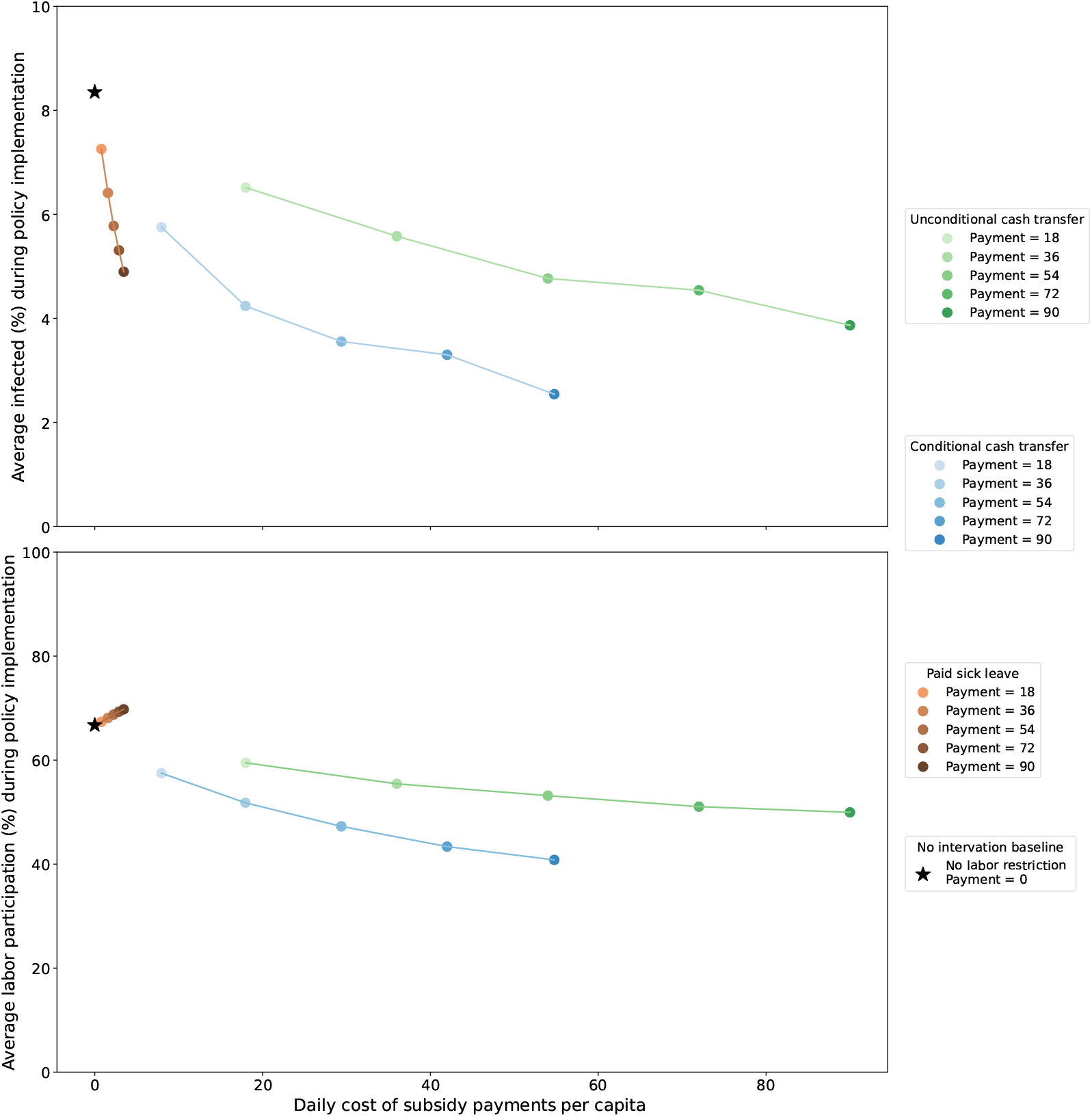
Average infection and labor supply by different policy interventions. This figure plots the average share of the population that is infected and working under different policy interventions as a function of their income loss. Panel a plots the average infection rates over the time period when the policies are in effect, while panel b plots the average share of the population that chose to work during this time period. The cost captures government spending to fund these cash transfers. The simulated policies include: a labor restriction policy that limits how much of the population is able to choose to work; an unconditional cash transfer that is delivered to all individuals each period; a conditional cash transfer that is delivered to individuals that choose to not work each period; and a paid sick leave policy, which allows infected individuals to earn their full wage if they choose to not work while infected.

